# Real-World Effectiveness of a Third Dose of mRNA-1273 versus BNT162b2 on Inpatient and Medically Attended COVID-19 among Immunocompromised Adults in the United States

**DOI:** 10.1101/2024.01.30.24302015

**Authors:** Tianyu Sun, Linwei Li, Katherine Mues, Mihaela Georgieva, Brenna Kirk, James Mansi, Nicolas Van de Velde, Ekkehard Beck

**Affiliations:** Moderna, Inc., Cambridge, MA, USA; Aetion, Inc., New York, NY, USA

**Author notes:** Corresponding author: Tianyu Sun, PhD.

**Keywords:** COVID-19, immunocompromised, mRNA-1273, SARS-CoV-2, vaccine effectiveness

## Abstract

Recent data have shown elevated infection rates in several subpopulations at risk of SARS-CoV-2 infection and COVID-19, including immunocompromised (IC) individuals. Previous research suggests that IC persons have reduced risks of hospitalization and medically-attended COVID-19 with 2 doses of mRNA-1273 (SpikeVax; Moderna) compared to two doses of BNT162b2 (Comirnaty; Pfizer/BioNTech). The main objective of this retrospective cohort study was to compare real-world effectiveness of third doses of mRNA-1273 versus BNT162b2 at multiple time points on occurrence of COVID-19 hospitalization and medically-attended COVID-19 among IC adults in the US. The HealthVerity (HV) medical and pharmacy claims database, which contains data from >330 million patients, was the data source. Both subgroup and sensitivity analyses were conducted in addition to the core comparisons noted. In propensity score-adjusted analyses, receiving mRNA-1273 vs BNT162b2 as third dose was associated with 32% (relative risk [RR] 0.68; 95% confidence interval [CI] 0.51-0.89), 29% (0.71; 0.57-0.86), and 23% (0.77; 0.62-0.93) lower risk of COVID-19 hospitalization after 90, 180, and 270 days, respectively. Corresponding reductions in medically-attended COVID-19 were 8% (0.92; 0.86-0.98), 6% (0.94; 0.90-0.98), and 2% (0.98; 0.94-1.02), respectively. Our findings suggest a third dose of mRNA-1273 is more effective than a third dose of BNT162b2 in preventing COVID-19 hospitalization and breakthrough medically-attended COVID-19 among IC adults in the US.

## INTRODUCTION

By September 2023, the United States (US) had registered >6 million cumulative hospitalizations due to cases of severe acute respiratory syndrome coronavirus 2 (SARS-CoV-2) infection and novel coronavirus disease 2019 (COVID-19), and >1 million associated deaths [1]. Although the availability of two mRNA vaccines, mRNA-1273 (SpikeVax; Moderna) and BNT162b2 (Comirnaty; Pfizer/BioNTech), has proved to be a critical tool against SARS-CoV-2 infection and COVID-19, there is a paucity of real-world data on the long-term effectiveness of vaccination. Recent data have shown increasing rates of infection in certain subpopulations at elevated risk of COVID-19 related morbidity and mortality, including immunocompromised individuals [2].

The safety and efficacy of two doses of both mRNA-1273 and BNT162b2 in reducing the risk of SARS-CoV-2 infection and of severe outcomes from COVID-19 in the general population and in adolescents has been demonstrated in phase 3 clinical trials [3–5] and in real-world studies [6, 7]. These results subsequently have been replicated in further observational studies, providing continued real-world evidence of the protective effects of two doses of both mRNA vaccines against SARS-CoV-2 infection, COVID-19 related hospitalization, and in-hospital mortality, despite the emergence of variants [6, 8, 9].

However, although high vaccine efficacy (VE) has been reported for both mRNA vaccines in the general population, these trials excluded immunocompromised (IC) individuals, such as those with underlying immunocompromising conditions and those prescribed immune-modifying therapies [10, 11]. Subsequent real-world data in the vaccinated IC population receiving two doses showed attenuated VE, with a higher risk of infection, hospitalization, death, persistent infection and shedding, viral evolution, reduced antibody and neutralization titers, and infection of household contacts [12]. In addition, observational data have suggested that there is a progressive dose- and time-dependent reduction in protection against SARS-CoV-2 infection among persons receiving a primary series of two doses [13–15].

In view of these increased risks, third and fourth doses of mRNA-based vaccines have been investigated in this vulnerable group [16] and, accordingly, both mRNA-1273 (50 or 100 μg; individuals aged ≥18 years) and BNT162b2 (30 μg; individuals aged ≥12 years) have been authorized and recommended for administration of a third primary dose ≥1 month after completion of the primary series in moderately to severely IC individuals [17, 18].

Our previous study showed that IC persons had reduced risks of hospitalization and medically-attended COVID-19 with two doses of mRNA-1273 compared to two doses of BNT162b2 [19], and the present study is an update assessing the comparative effectiveness of a third dose of both mRNA vaccines, regardless of number of doses included in the primary series. A matched cohort study showed that three doses of mRNA-1273 were associated with a significantly higher relative vaccine effectiveness (rVE) against SARS-CoV-2 infection and severe COVID-19 related outcomes compared to two doses, not only across subgroups of demographic and clinical characteristics but also in immunocompromised individuals [20, 21], and other studies have suggested that a third dose of BNT162b2 is safe and effective in preventing severe COVID-19-related outcomes [22, 23].

Previous influenza vaccine trials have demonstrated that high dose vaccines lead to improved immune responses in IC individuals compared with standard dose vaccines, which suggests that a high dose vaccine offers greater effectiveness for IC populations [24]. Although both mRNA-1273 and BNT162b2 employ the same mode of action, the composition of each vaccine is different, with the mRNA dosage and type of lipid nanoparticles used in the delivery system differing between vaccines. Also, the mRNA-1273 primary series contained 100 μg of mRNA vaccine and 50 μg for the booster [17], whereas BNT162b2 contained 30 μg of mRNA vaccine for each primary and booster dose [18]. Observational studies have shown differences between the two mRNA vaccines both in terms of immune response [25] and clinical effectiveness [19] in IC populations [24].

### Study objective

The primary objective of this study was to compare real-world effectiveness of a third dose of mRNA-1273 versus a third dose of BNT162b2 at multiple time points (90, 180, and 270 days after start of follow-up) on occurrence of COVID-19 hospitalization and of medically-attended breakthrough COVID-19 among adults in the US who are immunocompromised and have received two doses of mRNA-based vaccine.

## METHODS

### Study design

This was a retrospective cohort study designed to compare real-world effectiveness of a third dose of mRNA-1273 versus a third dose of BNT162b2 on COVID-19 hospitalization and medically-attended breakthrough COVID-19 among US adults who are immunocompromised and have already received two doses of mRNA-based COVID-19 vaccines. The data source for this study was the HealthVerity (HV) aggregated and anonymized medical and pharmacy claims database, which holds data from >150 US healthcare payers for >330 million patients. Claims were drawn from the time period December 11, 2020 through August 31, 2022, a period which saw the emergence of dominant SARS-CoV-2 variants [26]. All available data of eligible patients were used in the derivation of baseline variables which were not otherwise defined or specified. At the time of data abstraction, the HV database contained approximately 3 million individuals with a third dose of mRNA-1273 and 4 million individuals with a third dose of BNT162b2, identified in HV closed-claim data feeds.

The sample of patients identified from closed claims (specifically private source 20) was enriched with an additional claims data feed (private source 17) linked through HealthVerity. This additional data feed includes adjudicated pharmacy claims sourced from a pharmacy benefit manager (PBM) and an associated pharmacy enrollment file. For a proportion (approximately 23%) of patients in private source 17, adjudicated medical claims are linked to the pharmacy enrollment file. For patients with linked medical claims in private source 17, 100% of adjudicated medical claims (i.e., all claims for those patients) are observable over the period in which a patient is enrolled in the PBM plan.

The index date (day 0) was defined as the day when study participants received a third dose of COVID-19 vaccine, either BNT162b2 or mRNA-1273. The baseline period was defined as the one-year period prior to index date. The start of follow-up was defined as 14 days after the index date. Patients were followed until the earliest of (a) receiving any additional COVID-19 vaccine, (b) observed occurrence of study outcome(s), (c) the date of a break in continuous enrollment in a study participant’s health plan, or (d) August 31, 2022 (Figure 1).

**Figure 1.**
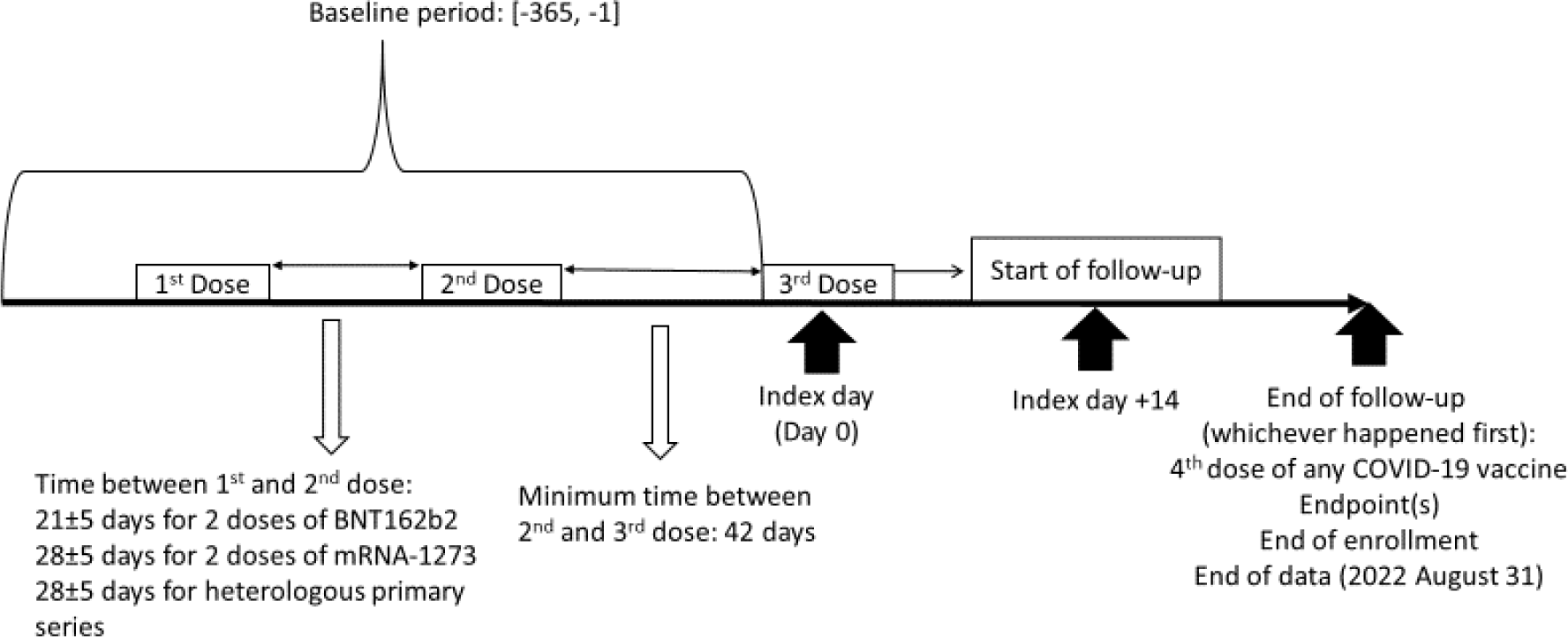
Study design schema.

### Study Population

The study population included IC adults in the US who had received three doses of mRNA-based COVID-19 vaccines (mRNA-1273 and BNT162b2) irrespective of the combination of the vaccines (e.g., three homologous mRNA-1273 or BNT162b2, or a heterologous combination of these), with the third dose considered the vaccine dose of interest, and the date of that dose establishing the index date. Immunocompromised individuals also were required to have been continuously enrolled in both medical and pharmacy insurance for at least one year before receiving the third dose and identified as immunocompromised on or before receiving the third dose.

#### Inclusion criteria

Individuals were eligible for inclusion in the analysis if they were aged ≥ 18 years on the index date, had continuous enrollment in medical or pharmacy plans during a 365-day baseline period prior to and including index date (i.e., no gaps were allowed), and had evidence of having received three doses of mRNA-based COVID-19, the first dose being on or after December 11, 2020. The required window between the first and second dose was determined by the combination of the first two doses: 28 ± 5-days for homologous 2 doses of mRNA-1273, 21 ± 5-days for homologous two doses of BNT162b2, and 28 ± 5-days for heterologous two doses. The third dose must have been received at least 42 days after the second dose. In addition, individuals had to be identified as immunocompromised based on evidence of one or more of the following criteria [19]: (a) blood or stem cell transplant in the two years prior to the index date; (b) history of organ transplant and taking immunosuppressive therapy within the 60 days prior to index date; (c) active cancer treatment in the 180 days prior to index date with an active cancer diagnosis in the 365 days prior to treatment; (d) prior history of a primary immunodeficiency disorder (e.g., for conditions such as DiGeorge syndrome and Wiskott-Aldrich syndrome); (e) history of an HIV diagnosis code prior to index date; or (f) immunosuppressive therapy in the 60 days prior to the index date.

#### Exclusion criteria

Individuals were excluded from the analysis if there was any evidence of the following: (a) receiving a non-mRNA COVID-19 vaccine up to and including the index date; (b) receipt of >1 mRNA vaccine dose on day 0; (c) receipt of any COVID-19 vaccine during day 1 through day 13, inclusive; (d) COVID-19 or related infectious disease during day 0 through day 13, inclusive; (e) missing or unknown sex or age on the index date; or (f) missing or unknown geographic region.

### Exposure

The comparison cohort was defined as those individuals who received the mRNA-1273 vaccine as the third dose after completion of two primary doses of mRNA-based COVID-19 vaccines within the time window specified above. The third dose could be either 50 µg or 100 µg of vaccine, depending on whether it was administered as part of a three-dose primary series or as a two-dose primary series with 100 µg dosing followed by a 50 µg booster dose. The reference cohort was defined as individuals who received the BNT162b vaccine as their third dose after completing two primary mRNA-based COVID-19 vaccines. Vaccine administrations were identified using Current Procedural Terminology (CPT) and/or National Drug Code (NDC) codes, with time windows between doses based on health authority recommendation [27]. The list of relevant codes is provided in Supplemental Table 1. For the first two doses, only the primary series codes were used. As noted previously, the third mRNA based COVID-19 vaccine dose was allowed to be either part of a three-dose primary series (as recommended for the IC population) or a booster dose administered following a two-dose primary series.

### Outcomes

Breakthrough COVID-19 hospitalization was identified based on presence of an International Classification of Diseases, 10^th^ edition (ICD-10) diagnosis code for COVID-19 (U07.1) in any diagnosis field coincident with a hospitalization episode. HV does not include an inpatient indicator in medical claims for de-identification purposes. Inpatient claims were distinguished from outpatient claims by place of service, revenue code and bill type codes. The hospitalization episode was created by combining concurrent/adjacent inpatient claims with a 4-day allowable gap. To increase specificity, we further required evidence of respiratory distress during the same episode, such as bronchitis, cough, or use of supplemental oxygen [28,29]. The full list of diseases and codes are provided in Supplemental Table 2. Medically-attended breakthrough COVID-19 was identified using the same U07.1 ICD-10 diagnosis code in any place of service setting (i.e., inpatient, outpatient, emergency room, or urgent care).

### Baseline Covariates

In addition to age, sex, geographic region, insurance type, and calendar month of index date, the following healthcare resource utilization measures and comorbid conditions were identified as potential confounders through literature review: (a) number of prior hospitalizations; (b) number of outpatient visits during baseline period; (c) number of unique immunosuppressive therapies used during baseline period; (d) the Charlson Quan Comorbidity Score during baseline period [30]; (e) the Kim claims-based frailty score during baseline period [31]; (f) recent medically-attended breakthrough COVID-19 infection (within six months prior to the index date); and (g) prior history of comorbid conditions. A full list of conditions is presented in Table 1. The algorithm used to derive these variables from claims data was documented in supplemental material published by Mues et al. [19].

**Table 1.**
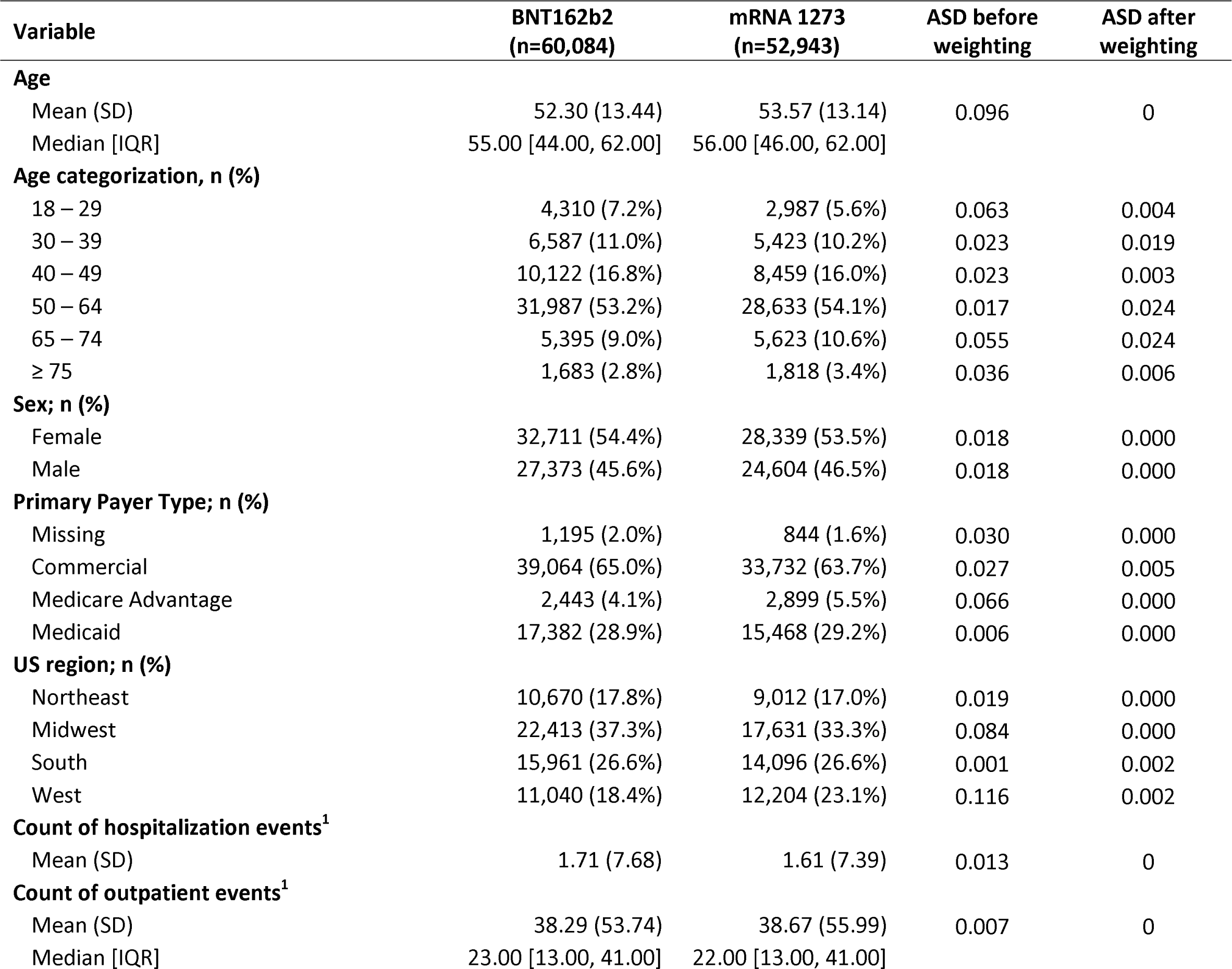
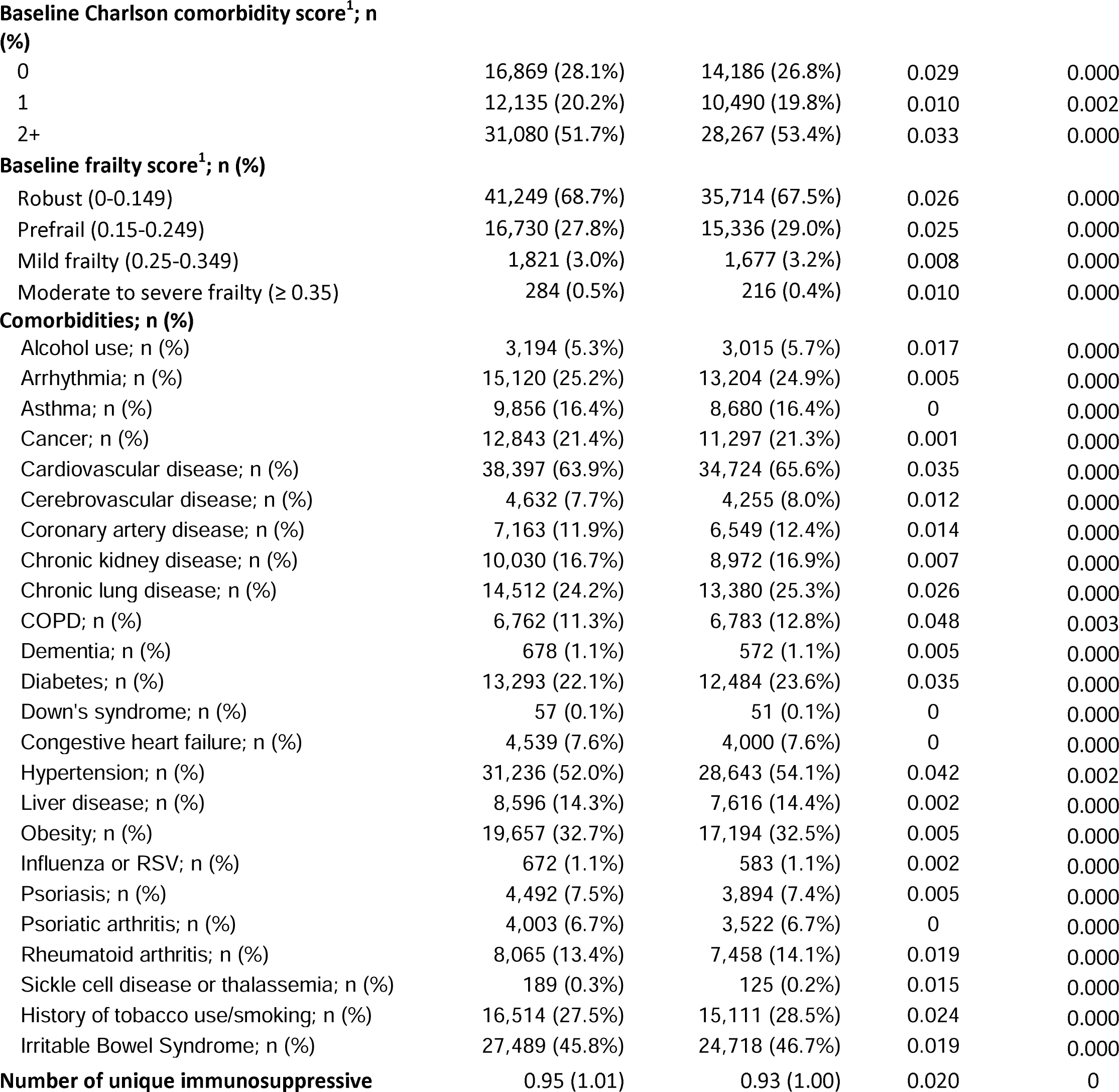
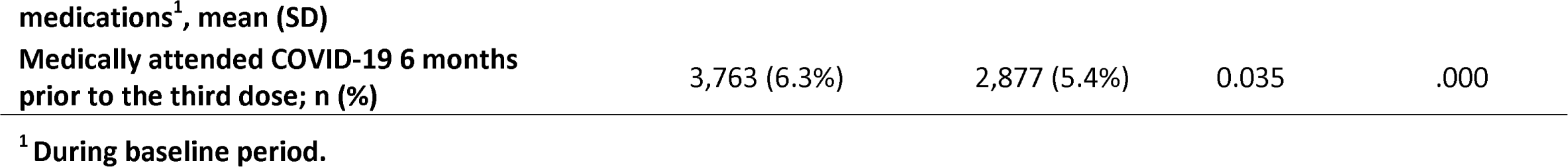
Baseline characteristics of immunocompromised adults with a third dose regimen of BNT162b2 or mRNA-1273.

### Statistical analysis

Distributions of baseline variables within each vaccine cohort were described as numbers and percentages for categorical variables, and as mean (standard deviation), and median (interquartile range) for continuous variables. Absolute standardized differences (ASDs) were calculated to determine the balance of baseline variables (ASD ≤0.10) between the two cohorts prior and post weighting [32, 33]. Crude incidence rates for the two endpoints were calculated for each vaccine cohort and reported on a per thousand person-year (TPY) basis with associated 95% confidence intervals (CI). Crude Kaplan-Meier curves with 95% CIs were plotted.

Propensity score (PS) based methods were employed to account for measured baseline confounders [32]. Logistic regression was used to calculate the PS. Baseline covariates that were hypothesized to be confounding variables were included. Since participants had a negligible percentage of missing baseline demographic data (age, sex, and geographic region), persons with missing data for one or more of these data elements were excluded without imputation (801 out of 6.42 million; Figure 2). The inverse probability of treatment weighting (IPTW) method was applied in the main analysis. The weights were calculated as 1/PS for the mRNA-1273 group and 1/(1-PS) for the BNT162b2 group. A Cox proportional hazard model was initially used to estimate hazard ratios comparing two cohorts on both endpoints. However, the proportional hazard assumption was not met. A non-parametric method was substituted, and crude and weighted Kaplan-Meier curves were generated to estimate event-free survival probabilities at multiple time points (90, 180 and, 270 days). The comparison then was conducted on both relative and absolute scales: relative risk (RR) and risk difference (RD) to compare event-free survival probabilities between two cohorts at multiple time points. The 95% CI was obtained by bootstrapping with 1000 replicates.

**Figure 2.**
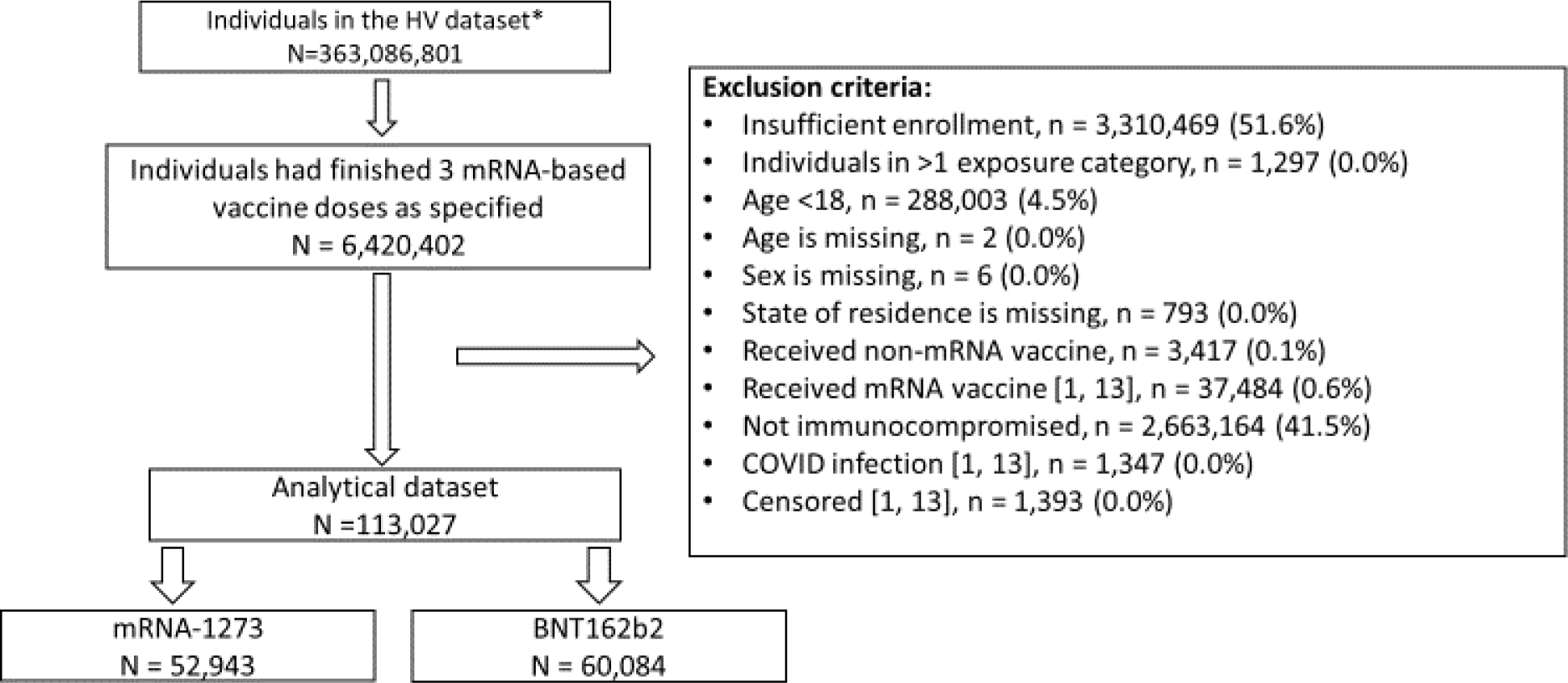
Participant attrition indicating the patient size before inverse probability of treatment weighting. * The entire HV dataset includes multiple sources of both closed and open claims during 2015 - 2022. Individuals from different sources and years can be counted duplicate and since many did not have enrollment file (such as open claims).

**Figure 3.**
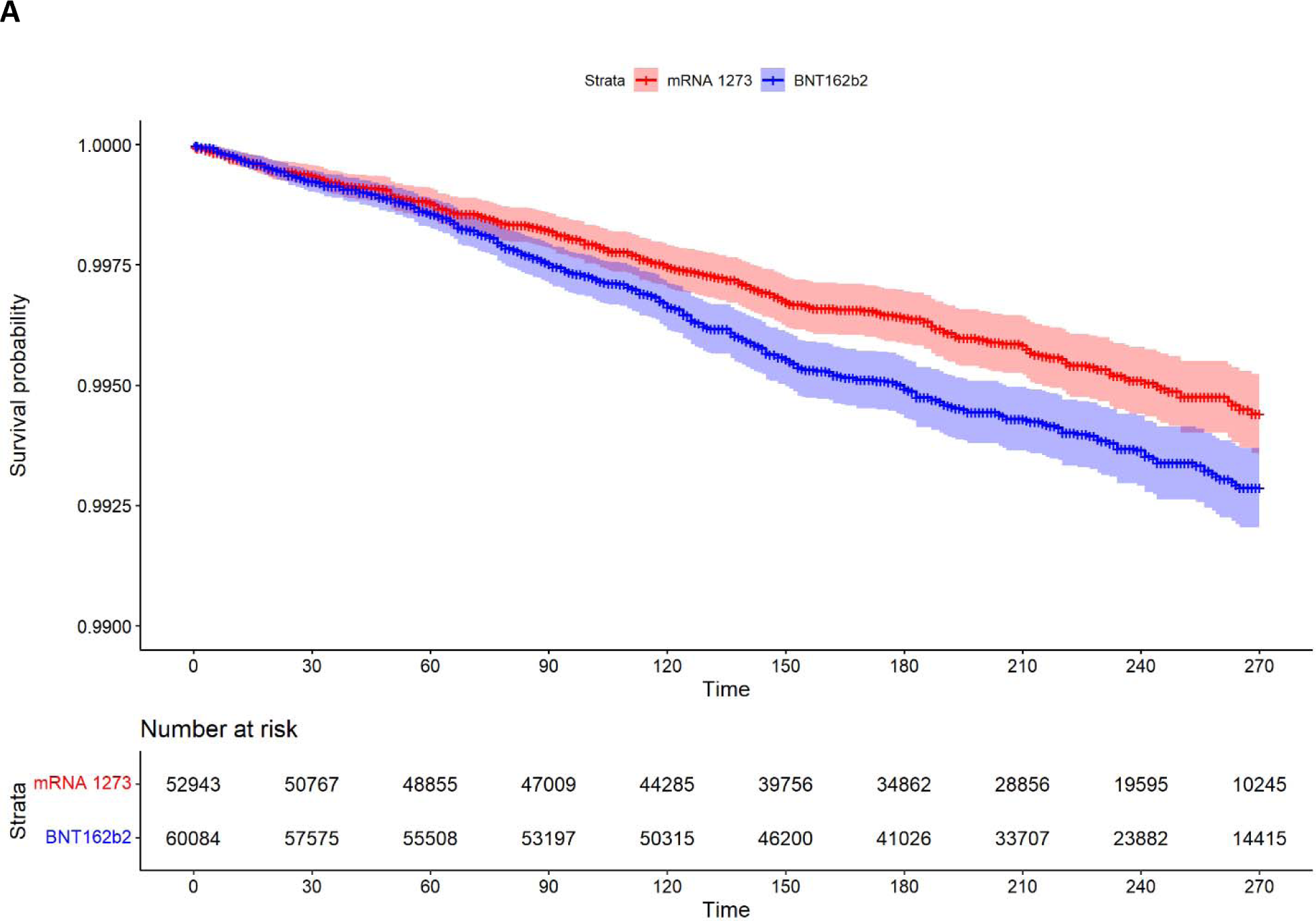
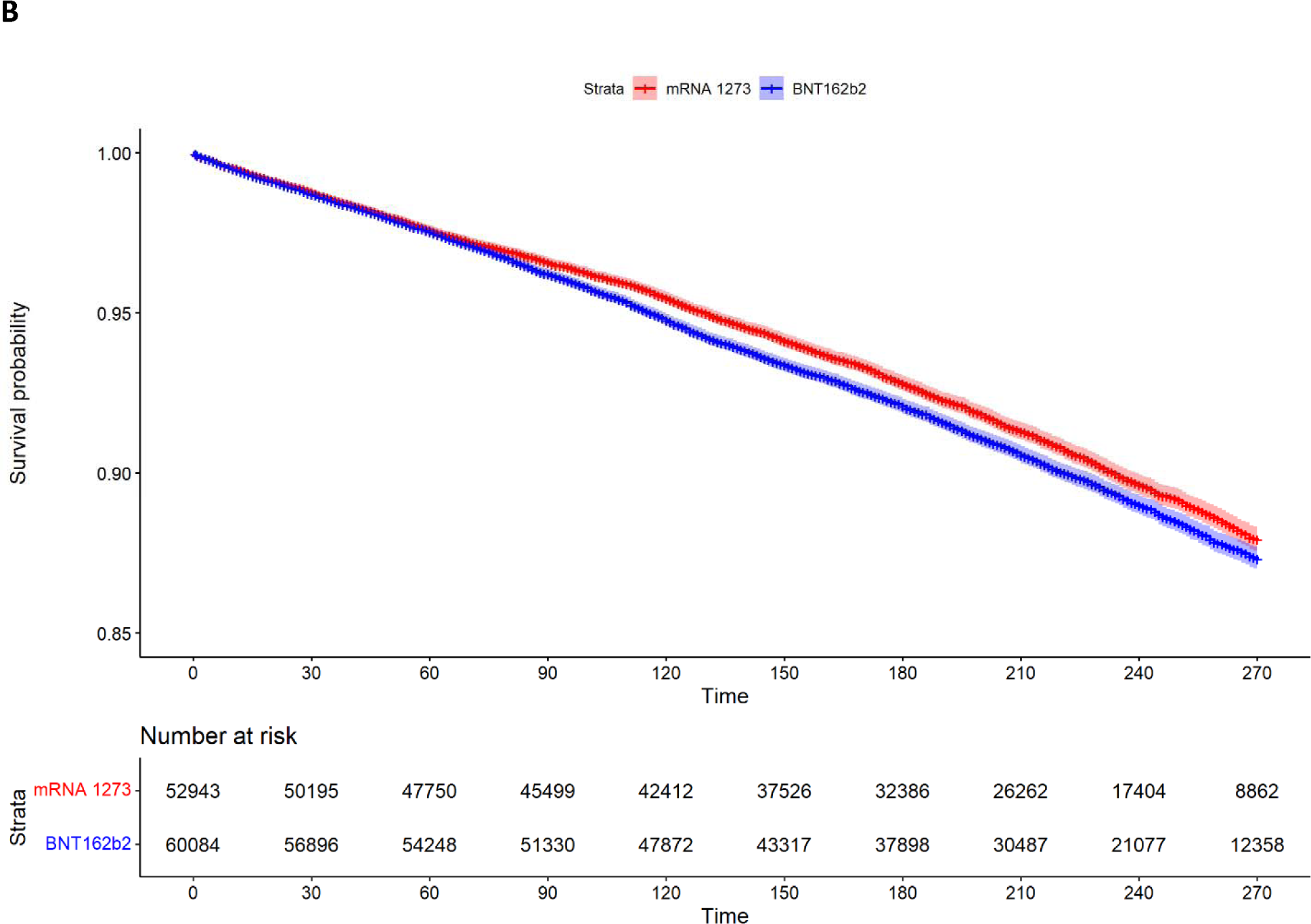
Kaplan–Meier plots of mRNA-1273 versus BNT162b cohort on A) COVID-19 hospitalization; B) Medically-attended COVID-19, by day 270.

### Subgroup analysis

Descriptive analyses were conducted for the following pre-specified subgroups of interest: participants aged 65+; participants without medically-attended COVID-19 in the 6 months prior to index day; participants with primary immunodeficiency; participants with active cancer; and participants diagnosed with HIV.

### Sensitivity Analysis

A series of sensitivity analyses were performed: 1) a more sensitive definition of COVID-19 hospitalization was tested, specifically requiring only a ICD-10 diagnosis code (U07.1) in an inpatient setting; 2) a second alternative definition of COVID-19 hospitalization in which a COVID-19 hospitalization was captured using more restrictive definition of respiratory distress (cf. Supplemental Table 2); 3) using both open and closed claims from HV for outcome identification purpose; and 4) repeating the analyses using PS matching with a nearest neighbor approach and caliper = 0.01.

### Software

Data cleaning and cohort selection was performed using the Aetion Evidence Platform: Substantiate software application for real-world data analysis, which has been scientifically validated for observational cohort studies using large healthcare databases [34]. All statistical analyses were conducted on SAS 9.4 (SAS Institute Inc.); plots were generated using R (4.3.1).

### Ethical considerations

The study was designed to facilitate an understanding of real-world clinical practice, and no tests, treatments, or investigations were conducted. Since the study utilized anonymized retrospective data, no identifiable protected health information was abstracted.

## RESULTS

A total of 113,027 immunocompromised adults were identified for inclusion in the primary analysis; of these, 52,943 and 60,084 received a third dose vaccination with mRNA-1273 and BNT162b2, respectively (Figure 2; Table 1). Individuals in the mRNA-1273 cohort were slightly older than in the BNT162b2 cohort (mean ages of 53.6 and 52.3 years, respectively) and more than one-half were aged 50-64 years. There were slightly fewer women in the mRNA-1273 cohort (54.4% vs. 53.5%, respectively; Table 1). Most patients were commercially insured (more than 60% among both cohorts). This immunocompromised population sample had a variety of comorbid conditions: for instance, in each cohort more than 60% had cardiovascular disease and more than one-half had hypertension. They also were actively seeking healthcare attention: on average, they had 38 outpatient claims during a year. In general, most demographics and comorbidities were comparable between these two cohorts except for calendar month of index date and geographic region. Since the BNT162b2 booster was approved earlier than mRNA-1273, the calendar month/year of index date tended to be earlier among individuals receiving BNT162b2 vs. mRNA-1273: 44.7% patients in the BNT162b2 cohort received their third dose during October-December 2021, while 45.8% patients in the mRNA-1273 cohort received their third dose during November-December 2021.

The median follow-up time of the two cohorts was comparable for both study endpoints (Table 2). The crude rate of COVID-19 hospitalization was lower among individuals who received a third dose of mRNA-1273 (7.89 per thousand person-years [TPY]; 95% CI, 6.88, 8.91) compared to those who received a third dose of BNT162b2 (9.99 per TPY; 95% CI, 8.93, 11.04). The crude rate of medically-attended breakthrough COVID-19 infection was also lower among the mRNA-1273 cohort (166.46 per TPY; 95% CI, 161.68, 171.25) compared to the BNT162b2 cohort (178.54 per TPY; 95% CI, 173.94, 183.15).

**Table 2.** Descriptive statistics of immunocompromised adults with a third dose of BNT162b2 or mRNA-1273 during follow-up.

|  | BNT162b2<br>N = 60,084 | mRNA 1273<br>N = 52,943 |
| --- | --- | --- |
| **Medically-attended COVID-19** |  |  |
| Total person-years | 32,395 | 27,970 |
| Number of events | 5,784 | 4,656 |
| Rate per 1,000 person-years | 178.54 | 166.46 |
| Median follow-up time (days) [IQR] | 211 [139, 258] | 209 [138, 253] |
| **COVID-19 hospitalization with respiratory distress** |  |  |
| Total person-years | 34,251 | 29,402 |
| Number of events | 342 | 232 |
| Rate per 1,000 person-years | 9.99 | 7.89 |
| Median follow-up time (days) [IQR] | 220 [157, 267] | 217 [150, 258] |

In the main comparison analyses, after IPTW, the measured baseline variables were considered balanced between the two cohorts (ASD ≤0.10) (Table 1). The weighted RR showed that, compared to BNT162b2, receiving mRNA-1273 as the third dose was associated with 32% (RR = 0.68, 95% CI: 0.51, 0.89), 29% (RR= 0.71, 95% CI: 0.57, 0.86), and 23% (RR= 0.77, 95% CI: 0.63, 0.93) lower risk of COVID-19 hospitalization after 90, 180, and 270 days, respectively (Table 3). The associations were statistically significant at all three time points. For medically-attended COVID-19, the magnitude of association was lower but consistent: receiving mRNA-1273 as the third dose was associated with 8% (RR= 0.92, 95% CI: 0.86, 0.98), 6% (RR= 0.94, 95% CI: 0.90, 0.98), and 2% (RR= 0.98, 95% CI: 0.94, 1.02) lower risk of medically-attended breakthrough COVID-19 after 90, 180, and 270 days. The association was statistically significant at 90 and 180 days, but not at 270 days. The absolute scale showed similar results: compared to BNT162b, receiving mRNA-1273 as the third dose was associated with 0.8, 1.5, and 1.7 fewer COVID-19 hospitalizations per thousand patients vaccinated after 90, 180, and 270 days, and 3.1, 5.0, and 3.0 fewer medically-attended breakthrough COVID-19 cases per thousand patients vaccinated after 90, 180, and 270 days (Table 3).

**Table 3.**
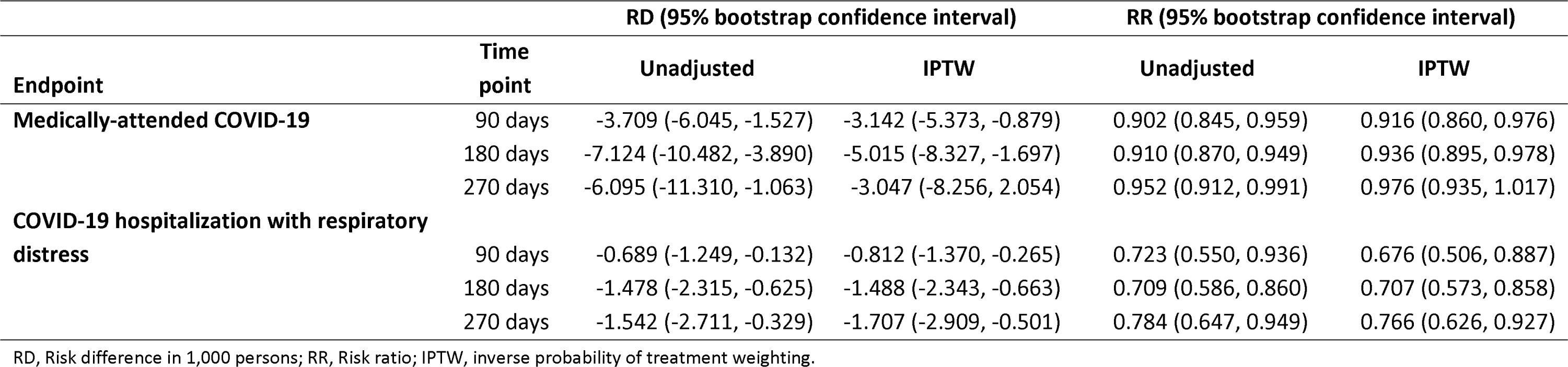
Comparative effectiveness of third dose of mRNA-1273 BNT162b2 on study endpoints among immunocompromised adults.

| Endpoint | Time point | RD (95% bootstrap confidence interval) |  | RR (95% bootstrap confidence interval) |  |
| --- | --- | --- | --- | --- | --- |
|  |  | Unadjusted | IPTW | Unadjusted | IPTW |
| **Medically-attended COVID-19** | 90 days | -3.709 (-6.045, -1.527) | -3.142 (-5.373, -0.879) | 0.902 (0.845, 0.959) | 0.916 (0.860, 0.976) |
|  | 180 days | -7.124 (-10.482, -3.890) | -5.015 (-8.327, -1.697) | 0.910 (0.870, 0.949) | 0.936 (0.895, 0.978) |
|  | 270 days | -6.095 (-11.310, -1.063) | -3.047 (-8.256, 2.054) | 0.952 (0.912, 0.991) | 0.976 (0.935, 1.017) |
| **COVID-19 hospitalization with respiratory distress** | 90 days | -0.689 (-1.249, -0.132) | -0.812 (-1.370, -0.265) | 0.723 (0.550, 0.936) | 0.676 (0.506, 0.887) |
|  | 180 days | -1.478 (-2.315, -0.625) | -1.488 (-2.343, -0.663) | 0.709 (0.586, 0.860) | 0.707 (0.573, 0.858) |
|  | 270 days | -1.542 (-2.711, -0.329) | -1.707 (-2.909, -0.501) | 0.784 (0.647, 0.949) | 0.766 (0.626, 0.927) |

RD, Risk difference in 1,000 persons; RR, Risk ratio; IPTW, inverse probability of treatment weighting.

Results from sensitivity analyses provided further confidence to the main results. Findings were robust in response to differing definitions of COVID-19 hospitalization: adjusted RRs and RDs were comparable to the main analyses at all three different timepoints using either more sensitive or more specific definitions of COVID-19 hospitalization. Including open claims for outcome identification also provided similar results as the main analyses for both endpoints (Supplemental Table 3). We also tested the sensitivity of results to use of different analytical methods. All analyses, including the alternative endpoint definitions, were repeated using PS matching instead of IPTW. All measured baseline variables were balanced between cohorts after matching (Supplemental Table 4). The RRs and RDs were very similar between these methods on different endpoints at the three time points (Supplemental Tables 3 & 5).

Results also were similar among all five pre-specified subgroups: elderly adults (age 65+), three groups with important immunocompromising conditions (primary immunodeficiency, active cancer, and HIV), and a COVID-19 naïve group (i.e., persons with no evidence of medically-attended COVID-19 within 6 months prior to the index date), with the rate of COVID-19 hospitalization and medically-attended COVID-19 numerically lower in the mRNA-1273 cohort than in the BNT162b2 cohort (Supplemental Table 6). The statistical comparison was not conducted due to limited sample size and statistical power.

## DISCUSSION

Observational studies have shown that a third dose of the mRNA-1273 or BNT162b2 vaccines is associated with a reduced risk of SARS-CoV-2 infection and associated severe COVID-19 related outcomes in immunocompetent adults [9, 20, 21]. In this large retrospective cohort study, we examined the real-world effectiveness of a third dose of mRNA-1273 versus a third dose of BNT162b2 in IC adults. It extended the work of a previous study that compared the effectiveness of two doses of mRNA-1273 and BNT162b2 vaccines among IC adults in the US [19]. This previous study showed that receiving two doses of mRNA-1273 was associated with lower medically-attended breakthrough COVID-19, and breakthrough COVID-19 hospitalization compared to receiving two doses of BNT162b2 among a population of IC adults.

Our findings are consistent with the previously established evidence of real-world effectiveness of both the mRNA-1273 and BNT162b2 vaccines among immunocompromised individuals compared to unvaccinated or partially vaccinated individuals [35]. The results suggested that rates of medically-attended COVID-19 and of COVID-19 hospitalization were lower among individuals who received a third dose of mRNA-1273 than those who received a third dose of BNT162b2. The evidence that a third dose of the mRNA-1273 vaccine is effective in preventing medically-attended COVID-19 outcomes and subsequent hospitalizations in IC individuals also supports recommendations from other studies [12, 15].

The results reported here have strength and uniqueness compared with prior studies that can provide valuable research insights in this fast-evolving COVID-19 field. To reflect the practice of mixed vaccination, this study did not require a homologous vaccine series as inclusion criteria. Furthermore, the third vaccine dose was inclusive of two common regimens: a three-dose primary series as recommended for immunocompromised patients, and a two-dose primary series plus booster. This generates a study sample reflective of a broader exposed population and may mimic more closely the real-world scenario that has evolved during the pandemic. Also, the methodology we used allows exploration of the effect of rVE on endpoints at different time points during follow-up. We investigated rVE at 90, 180, and 270 days after start of follow-up and results showed that mRNA1273 was more effective than BNT162b2 across different durations of follow-up.

From a statistical methodology standpoint, the results would confer causal interpretation if all three causal inference assumptions are met: consistency, exchangeability, and positivity [36]. We identified important potential confounders from published literature, derived them from the claims data based on established algorithms, and implemented two PS-based methods to account for measured ones. Even before adjustment, most measured baseline variables were comparable between cohorts and all of them were considered balanced after weighting/matching (Table 1). We empirically assessed loss to follow-up between two comparison cohorts to identify potential selection bias. The censoring reasons were distributed similarly between cohorts (Supplemental Table 7), and we found no evidence of differential loss to follow-up which potentially might lead to selection bias. Comparative effectiveness was estimated by a non-parametric method. Although less commonly used than Cox proportional hazard regression, it has fewer underlying assumptions and can estimate RDs and RRs which have more straightforward interpretation then the hazard ratio [37]. PS-based methods could be easily incorporated and has been applied in similar studies [22, 23, 38].

There are several limitations of this study. First, due to the lack of randomization inherent in claims data, there might be residual confounding due to unmeasured confounders, such as lifestyle or occupation among others. Second, all variables, including exposure, outcomes, and baseline covariates, were derived from medical and/or pharmacy claims. Random errors in these claims could lead to measurement error. However, it is believed to be non-differential between cohorts and would only bias results towards the null [39]. To mitigate this risk, this study employed a COVID-19 ICD-10 code to identify claims associated with COVID-19; this code has been shown to have high sensitivity, specificity, positive predictive value, and negative predictive value in various data sources [40, 41, 42]. We further conducted a series of sensitivity analyses using more sensitive or more specific outcome definitions which yielded consistent results. Lastly, there were a few limitations of the data sources we employed. To protect the privacy of the participants, during the anonymization process, people who were aged 85 years or older were coded as age 94 years. We believe the impact of this systematic misclassification was negligible since the prevalence of this age group in the overall sample was minimal. The HV database captured mainly commercially insured patients with some Medicaid and Medicare Advantage enrollees. Extrapolation of the conclusion to other populations, such as the uninsured, general Medicaid or Medicare fee-for-service enrollees, should be undertaken with caution.

Despite such limitations, real-world studies such as that reported herein play an important part in highlighting areas of concern not addressed in clinical trials and provide evidence of the comparative effectiveness of interventions in individuals seen in routine clinical practice.

## CONCLUSION

The results from this observational comparative VE database study provide evidence that a third dose of mRNA-1273 is more effective than a third dose of BNT162b2 in preventing breakthrough medically-attended COVID-19 and subsequent hospitalizations among IC adults in the US.

## Data Availability

Individual-level data reported in this study are not publicly shared. Upon request, and subject to review, Aetion, Inc. may provide the de-identified aggregate-level data that support the findings of this study. De-identified data (including participant data as applicable) may be shared upon approval of an analysis proposal and a signed data access agreement.

## ACKNOWLEDGEMENTS AND DISCLOSURES

Medical writing and editorial assistance were provided by K Ian Johnson BSc, MBPS, SRPharmS of MEDiSTRAVA, in accordance with Good Publication Practice (GPP) guidelines, funded by Moderna, Inc., and under the direction of the authors.

## AUTHOR CONTRIBUTIONS

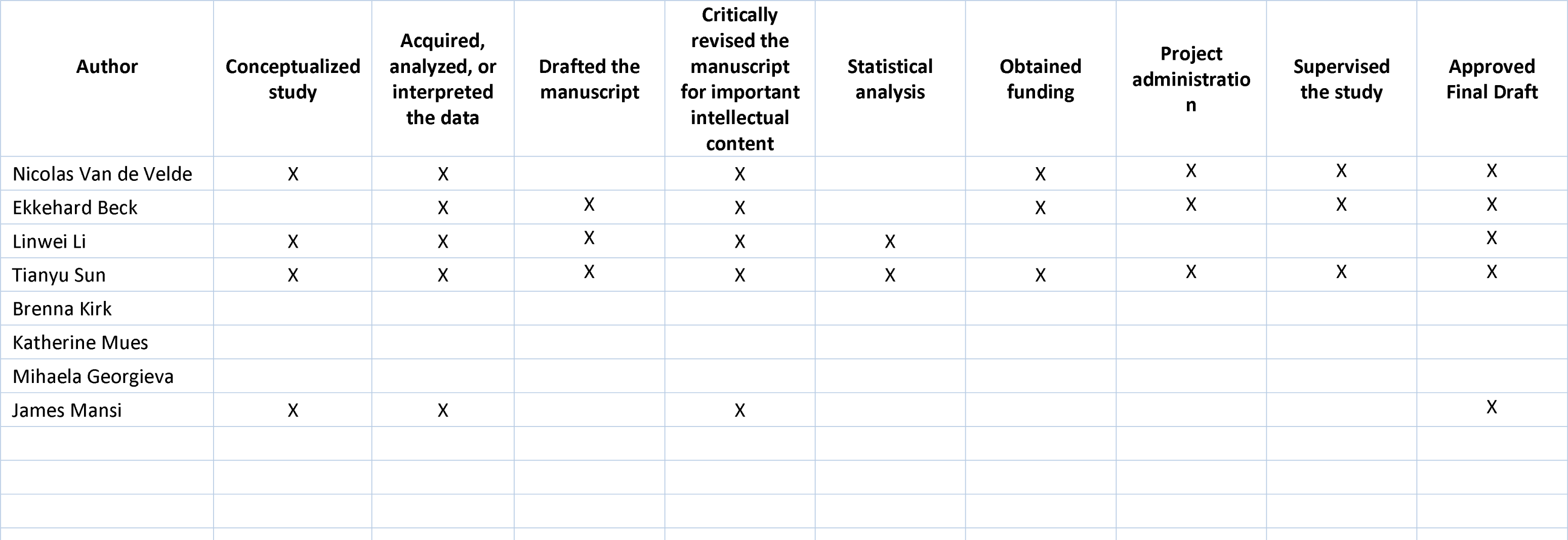

---

**Supplemental Table 1.** Codes to identify mRNA-1273 and BNT162b vaccines.

| mRNA-1273 primary series vaccine | mRNA-1273 booster vaccine | BNT-162b2 primary series vaccine | BNT-162b2 booster vaccine |
| --- | --- | --- | --- |
| CPT | CPT | CPT | CPT |
| 0011A | 91306 | 91300 | 0004A |
| 0012A | 0064A | 0001A | 0054A |
| 91301 | 0094A | 0002A | 91312 |
| **National Drug Code** | 91309 | 91305 | 0124A |
| 80777027310 | 91313 | 0051A | 0003A |
| 80777027315 | 0134A | 0052A | 0053A |
| 80777027398 | 0013A | **National Drug Code** | **National Drug Code** |
| 80777027399 | **National Drug Code** | 59267100001 | 59267030401 |
| 8077727315 | 8077727505 | 59267100002 | 5926703041 |
| 8077727398 | 80777027505 | 59267100003 | 5926703042 |
| 8077727399 | 8077728205 | 5926710001 | 59267030402 |
| 8077727310 | 80777028205 | 5926710002 | 59267140401 |
| 8077710011 | 8077728299 | 5926710003 | 5926714041 |
| 8077701001 | 80777028005 | 59267102501 | 5926714042 |
| 8077710099 | 80777028099 | 5926710251 | 59267140402 |
| 80777010011 | 80777028299 | 00069202501 |  |
| 80777010099 | 80777027599 | 0006920251 |  |
|  | 8077727599 | 00069202510 |  |
|  |  | 0069202501 |  |
|  |  | 0069202510 |  |
|  |  | 0069202525 |  |
|  |  | 00069202525 |  |
|  |  | 5926710252 |  |
|  |  | 59267102502 |  |
|  |  | 5926710253 |  |
|  |  | 59267102503 |  |
|  |  | 5926710254 |  |
|  |  | 59267102504 |  |

**Supplemental Table 2.**
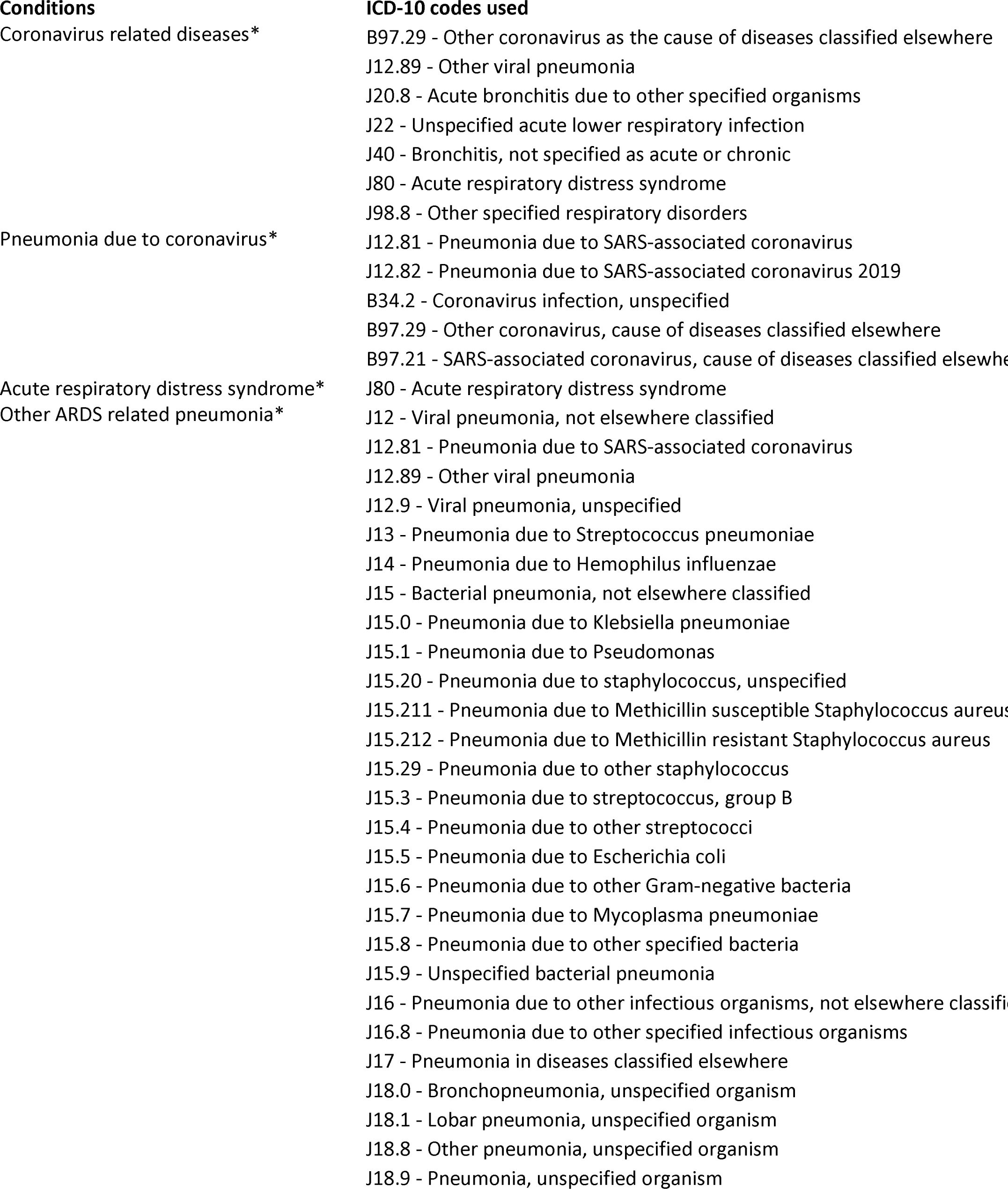
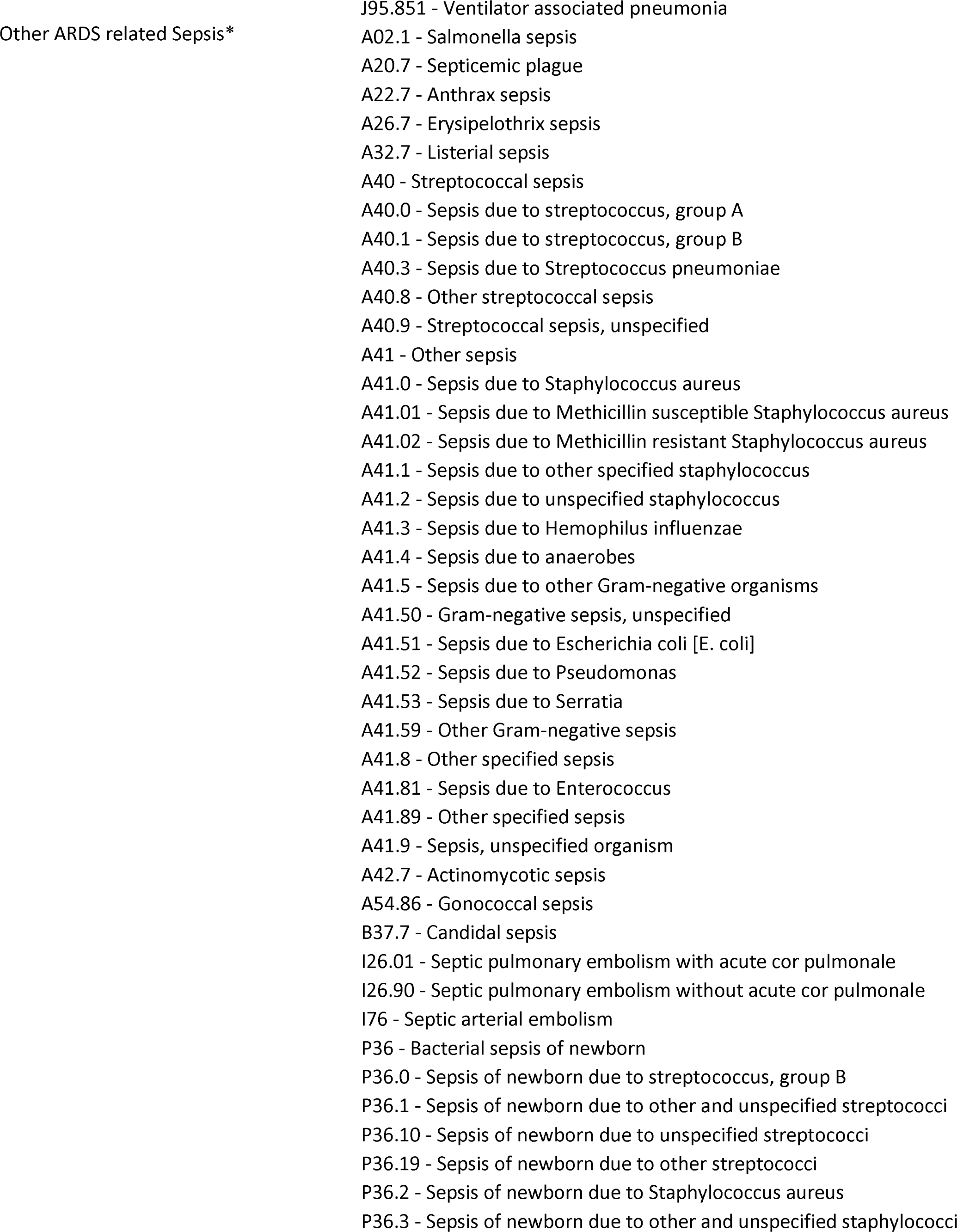
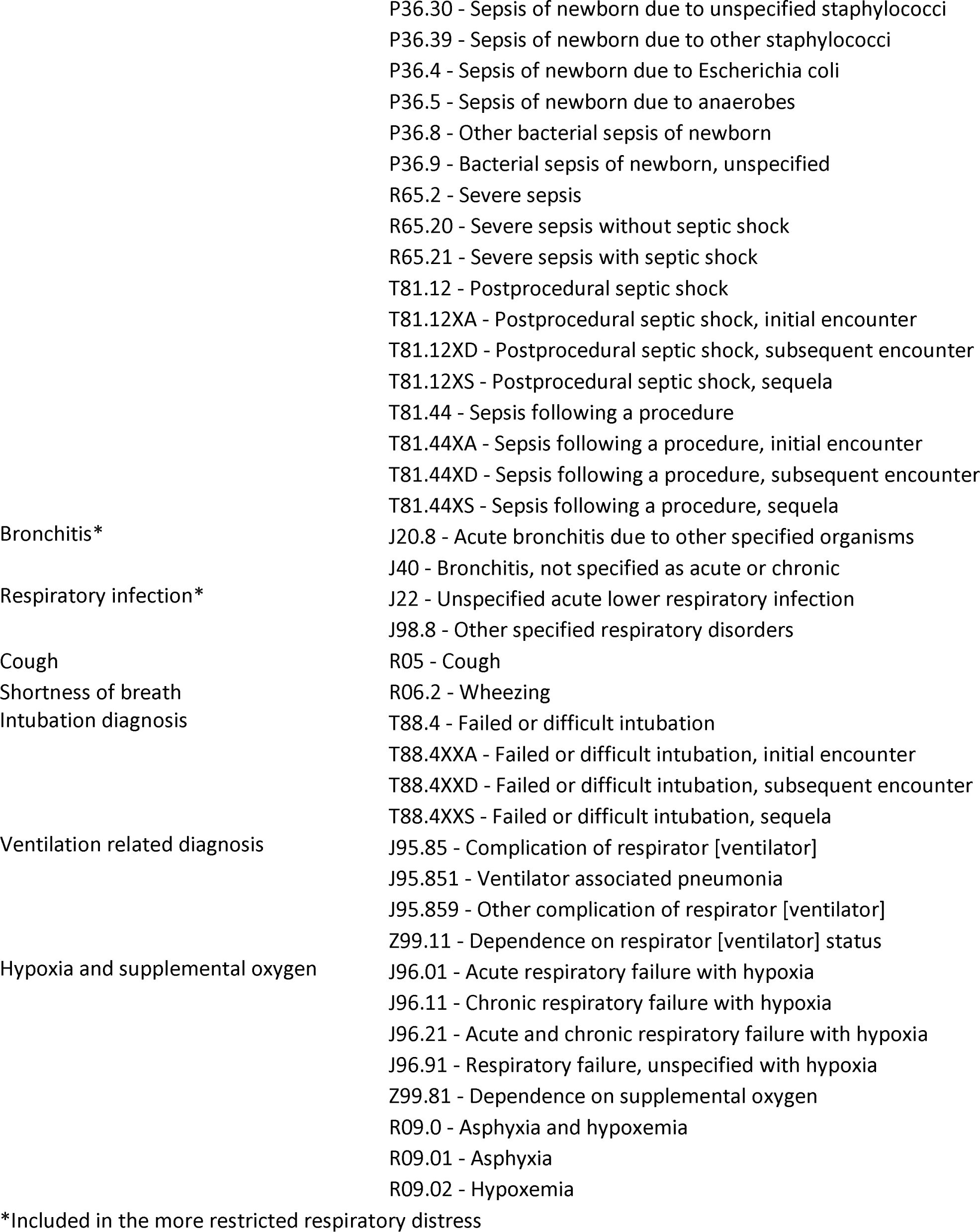
Diagnosis codes used to identify respiratory distress.

**Supplemental Table 3.** Sensitivity analyses results using different endpoint definitions and using both IPTW and PS-matching.

| Endpoint | Time point | RD (95% CI) |  |  | RR (95% CI) |  |  |
| --- | --- | --- | --- | --- | --- | --- | --- |
|  |  | Unadjusted | IPTW | PS matching | Unadjusted | IPTW | PS matching |
| COVID-19 hospitalization | 90 days | -0.974 (-1.627, -0.354) | -1.120 (-1.728, -0.507) | -1.202 (-1.879, -0.528) | 0.699 (0.538, 0.874) | 0.659 (0.515, 0.830) | 0.651 (0.506, 0.830) |
|  | 180 days | -1.695 (-2.760, -0.716) | -1.691 (-2.629, -0.714) | -1.768 (-2.755, -0.730) | 0.741 (0.613, 0.883) | 0.742 (0.621, 0.883) | 0.736 (0.619, 0.875) |
|  | 270 days | -2.337 (-3.726, -0.944) | -2.435 (-3.783, -1.088) | -2.491 (-4.021, -1.011) | 0.764 (0.649, 0.897) | 0.758 (0.646, 0.885) | 0.754 (0.640, 0.892) |
| COVID-19 hospitalization with more restricted respiratory distress | 90 days | -1.202 (-1.932, -0.489) | -1.202 (-1.694, -0.686) | -1.202 (-1.932, -0.489) | 0.651 (0.499, 0.838) | 0.651 (0.539, 0.779) | 0.651 (0.499, 0.838) |
|  | 180 days | -1.768 (-2.792, -0.785) | -1.768 (-2.506, -1.027) | -1.768 (-2.792, -0.785) | 0.736 (0.618, 0.872) | 0.736 (0.646, 0.835) | 0.736 (0.618, 0.872) |
|  | 270 days | -2.491 (-3.942, -1.030) | -2.491 (-3.661, -1.346) | -2.491 (-3.942, -1.030) | 0.754 (0.637, 0.889) | 0.754 (0.660, 0.863) | 0.754 (0.637, 0.889) |
| COVID-19 hospitalization with respiratory distress using both open and closed claims | 90 days | -0.758 (-1.300, -0.229) | -0.889 (-1.458, -0.292) | -0.942 (-1.609, -0.288) | 0.706 (0.546, 0.900) | 0.658 (0.499, 0.873) | 0.654 (0.486, 0.878) |
|  | 180 days | -1.547 (-2.379, -0.752) | -1.578 (-2.480, -0.773) | -1.762 (-2.809, -0.797) | 0.705 (0.582, 0.845) | 0.699 (0.572, 0.840) | 0.678 (0.540, 0.842) |
|  | 270 days | -1.763 (-2.922, -0.604) | -1.948 (-3.236, -0.725) | -2.040 (-3.334, -0.695) | 0.764 (0.637, 0.912) | 0.745 (0.614, 0.898) | 0.738 (0.605, 0.904) |
| Medically attended COVID-19 using both open and closed claim | 90 days | -3.653 (-5.929, -1.501) | -3.151 (-5.568, -0.789) | -2.709 (-5.112, -0.313) | 0.905 (0.851, 0.960) | 0.917 (0.859, 0.979) | 0.928 (0.869, 0.992) |
|  | 180 days | -7.031 (-10.417, -3.665) | -4.956 (-8.582, -1.350) | -5.346 (-8.842, -1.892) | 0.912 (0.872, 0.953) | 0.937 (0.894, 0.983) | 0.933 (0.890, 0.976) |
|  | 270 days | -5.817 (-10.917, -0.516) | -2.782 (-7.928, 2.869) | -3.696 (-9.461, 1.777) | 0.955 (0.916, 0.996) | 0.978 (0.939, 1.023) | 0.971 (0.927, 1.014) |

**Supplemental Table 4.** Baseline characteristics of mRNA-1273 and BNT1262b cohorts after propensity score matching.

|  | **BNT162b** | **mRNA 1273** | **ASD after matching** |
| --- | --- | --- | --- |
| Number of patients | 48747 | 48747 |  |
| **Month/Year of Cohort Entry Date** |  |  |  |
| February 2021; n (%) | 0 (0.0%) | 0 (0.0%) | 0 |
| March 2021; n (%) | 13 (0.0%) | 14 (0.0%) | 0.001 |
| April 2021; n (%) | 55 (0.1%) | 53 (0.1%) | 0.001 |
| May 2021; n (%) | 56 (0.1%) | 55 (0.1%) | 0.001 |
| June 2021; n (%) | 53 (0.1%) | 52 (0.1%) | 0.001 |
| July 2021; n (%) | 59 (0.1%) | 52 (0.1%) | 0.004 |
| August 2021; n (%) | 4,704 (9.6%) | 4,692 (9.6%) | 0.001 |
| September 2021; n (%) | 3,475 (7.1%) | 3,447 (7.1%) | 0.002 |
| October 2021; n (%) | 4,925 (10.1%) | 4,848 (9.9%) | 0.005 |
| November 2021; n (%) | 9,941 (20.4%) | 9,927 (20.4%) | 0.001 |
| December 2021; n (%) | 10,875 (22.3%) | 10,905 (22.4%) | 0.001 |
| January 2022; n (%) | 6,393 (13.1%) | 6,411 (13.2%) | 0.001 |
| February 2022; n (%) | 2,049 (4.2%) | 2,102 (4.3%) | 0.005 |
| March 2022; n (%) | 1,305 (2.7%) | 1,320 (2.7%) | 0.002 |
| April 2022; n (%) | 1,676 (3.4%) | 1,671 (3.4%) | 0.001 |
| May 2022; n (%) | 1,307 (2.7%) | 1,272 (2.6%) | 0.004 |
| June 2022; n (%) | 804 (1.6%) | 832 (1.7%) | 0.004 |
| July 2022; n (%) | 756 (1.6%) | 773 (1.6%) | 0.003 |
| August 2022; n (%) | 301 (0.6%) | 321 (0.7%) | 0.005 |
| **Age** |  |  |  |
| mean (sd) | 52.94 (13.11) | 52.89 (13.04) | 0.004 |
| median [IQR] | 56.00 [45.00, 62.00] | 55.00 [45.00, 62.00] |  |
| **Age categorization (18-29, 30-39, 40-49, 50-64, 65-74, 75+)** |  |  |  |
| 18 - <30; n (%) | 3,031 (6.2%) | 2,965 (6.1%) | 0.006 |
| 30 - <40; n (%) | 4,986 (10.2%) | 5,288 (10.8%) | 0.02 |
| 40 - <50; n (%) | 8,058 (16.5%) | 8,114 (16.6%) | 0.003 |
| 50 - <65; n (%) | 26,756 (54.9%) | 26,241 (53.8%) | 0.021 |
| 65 - <75; n (%) | 4,473 (9.2%) | 4,772 (9.8%) | 0.021 |
| >= 75; n (%) | 1,443 (3.0%) | 1,367 (2.8%) | 0.009 |
| **Female/male recat** |  |  |  |
| Female; n (%) | 26,286 (53.9%) | 26,267 (53.9%) | 0.001 |
| Male; n (%) | 22,461 (46.1%) | 22,480 (46.1%) | 0.001 |
| **Primary Payer Type** |  |  |  |
| Commercial; n (%) | 31,576 (64.8%) | 31,482 (64.6%) | 0.004 |
| Medicare Advantage; n (%) | 2,184 (4.5%) | 2,148 (4.4%) | 0.004 |
| Medicaid; n (%) | 14,189 (29.1%) | 14,297 (29.3%) | 0.005 |
| missing; n (%) | 798 (1.6%) | 820 (1.7%) | 0.004 |
| **Region (State re-categorization)** |  |  |  |
| Northeast; n (%) | 8,537 (17.5%) | 8,575 (17.6%) | 0.002 |
| Midwest; n (%) | 16,960 (34.8%) | 16,980 (34.8%) | 0.001 |
| South; n (%) | 13,226 (27.1%) | 13,170 (27.0%) | 0.003 |
| West; n (%) | 10,024 (20.6%) | 10,022 (20.6%) | 0 |
| **Count of hospitalization events, 365 days** |  |  |  |
| mean (sd) | 1.64 (7.38) | 1.62 (7.54) | 0.003 |
| median [IQR] | 0.00 [0.00, 0.00] | 0.00 [0.00, 0.00] |  |
| **Count of outpatient events, 365 days** |  |  |  |
| mean (sd) | 38.41 (54.48) | 38.34 (55.32) | 0.001 |
| median [IQR] | 23.00 [13.00, 41.00] | 22.00 [13.00, 40.00] |  |
| **Charlson comorbidity score categories, 365 days** |  |  |  |
| 0; n (%) | 13,323 (27.3%) | 13,425 (27.5%) | 0.005 |
| 1; n (%) | 9,811 (20.1%) | 9,734 (20.0%) | 0.004 |
| 2+; n (%) | 25,613 (52.5%) | 25,588 (52.5%) | 0.001 |
| **Frailty score categories, 365 days** |  |  |  |
| robust (0-0.149); n (%) | 33,175 (68.1%) | 33,212 (68.1%) | 0.002 |
| prefrail (0.15-0.249); n (%) | 13,856 (28.4%) | 13,839 (28.4%) | 0.001 |
| mild frailty (0.25-0.349); n (%) | 1,514 (3.1%) | 1,488 (3.1%) | 0.003 |
| moderate to severe frailty ($\geq 0.35$) ; n (%) | 202 (0.4%) | 208 (0.4%) | 0.002 |
| **Baseline comorbidities; n (%)** |  |  |  |
| Alcohol use; n (%) | 2,700 (5.5%) | 2,721 (5.6%) | 0.002 |
| Arrhythmia; n (%) | 12,110 (24.8%) | 12,165 (25.0%) | 0.003 |
| Asthma; n (%) | 7,982 (16.4%) | 7,997 (16.4%) | 0.001 |
| Cancer; n (%) | 10,348 (21.2%) | 10,352 (21.2%) | 0 |
| Cardiovascular disease; n (%) | 31,526 (64.7%) | 31,497 (64.6%) | 0.001 |
| Cerebrovascular disease; n (%) | 3,824 (7.8%) | 3,842 (7.9%) | 0.001 |
| Coronary artery disease; n (%) | 5,866 (12.0%) | 5,909 (12.1%) | 0.003 |
| Chronic kidney disease; n (%) | 8,235 (16.9%) | 8,149 (16.7%) | 0.005 |
| Chronic lung disease; n (%) | 11,992 (24.6%) | 12,038 (24.7%) | 0.002 |
| COPD; n (%) | 5,819 (11.9%) | 5,866 (12.0%) | 0.003 |
| Dementia; n (%) | 509 (1.0%) | 522 (1.1%) | 0.003 |
| Diabetes; n (%) | 11,141 (22.9%) | 11,090 (22.8%) | 0.002 |
| Down's syndrome; n (%) | 50 (0.1%) | 47 (0.1%) | 0.002 |
| Congestive heart failure; n (%) | 3,672 (7.5%) | 3,638 (7.5%) | 0.003 |
| Hypertension; n (%) | 25,888 (53.1%) | 25,813 (53.0%) | 0.003 |
| Liver disease; n (%) | 7,009 (14.4%) | 7,039 (14.4%) | 0.002 |
| Obesity; n (%) | 15,873 (32.6%) | 15,853 (32.5%) | 0.001 |
| Influenza or RSV; n (%) | 543 (1.1%) | 535 (1.1%) | 0.002 |
| Psoriasis; n (%) | 3,663 (7.5%) | 3,616 (7.4%) | 0.004 |
| Psoriatic arthritis; n (%) | 3,294 (6.8%) | 3,254 (6.7%) | 0.003 |
| Rheumatoid arthritis; n (%) | 6,736 (13.8%) | 6,702 (13.7%) | 0.002 |
| Sickle cell disease or thalassemia; n (%) | 119 (0.2%) | 125 (0.3%) | 0.002 |
| History of tobacco use/smoking; n (%) | 13,711 (28.1%) | 13,719 (28.1%) | 0 |
| Irritable Bowel Syndrome; n (%) | 22,540 (46.2%) | 22,575 (46.3%) | 0.001 |
| **Number of unique immunosuppressive medication** |  |  |  |
| mean (sd) | 0.94 (1.00) | 0.94 (1.00) | 0 |
| median [IQR] | 1.00 [0.00, 1.00] | 1.00 [0.00, 1.00] |  |
| **COVID-19 infection [-180,-1]; n (%)** | 2,774 (5.7%) | 2,799 (5.7%) | 0.002 |

**Supplemental Table 5.**
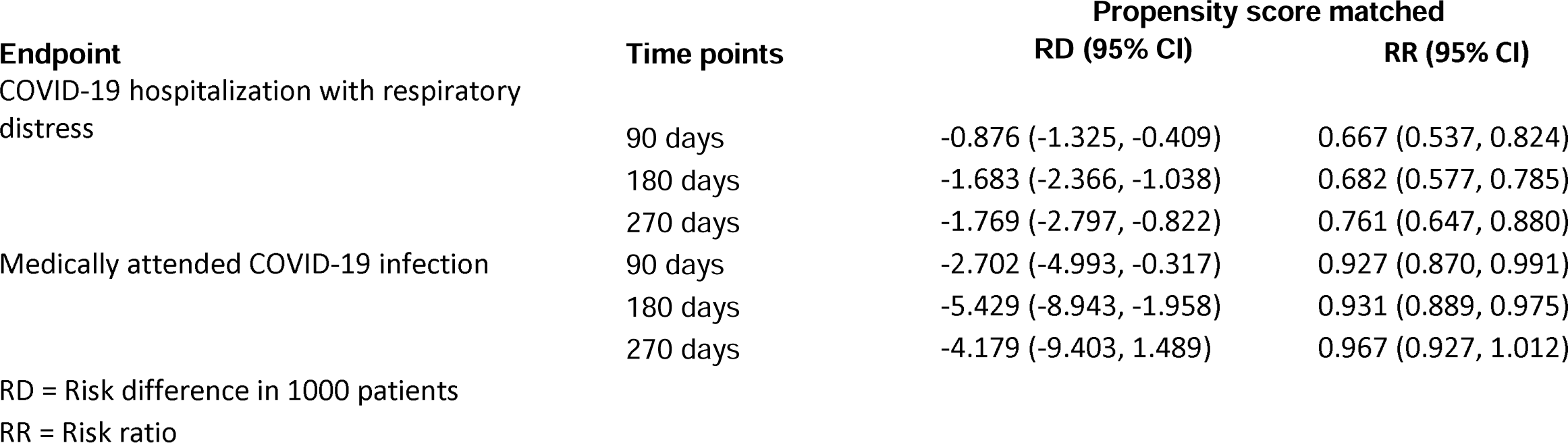
Sensitivity analysis results using PS matching.

| **Endpoint** | **Time points** | **Propensity score matched** |  |
| --- | --- | --- | --- |
|  |  | **RD (95% CI)** | **RR (95% CI)** |
| COVID-19 hospitalization with respiratory distress | 90 days | -0.876 (-1.325, -0.409) | 0.667 (0.537, 0.824) |
|  | 180 days | -1.683 (-2.366, -1.038) | 0.682 (0.577, 0.785) |
|  | 270 days | -1.769 (-2.797, -0.822) | 0.761 (0.647, 0.880) |
| Medically attended COVID-19 infection | 90 days | -2.702 (-4.993, -0.317) | 0.927 (0.870, 0.991) |
|  | 180 days | -5.429 (-8.943, -1.958) | 0.931 (0.889, 0.975) |
|  | 270 days | -4.179 (-9.403, 1.489) | 0.967 (0.927, 1.012) |

RD = Risk difference in 1000 patients
RR = Risk ratio

**Supplemental Table 6.**
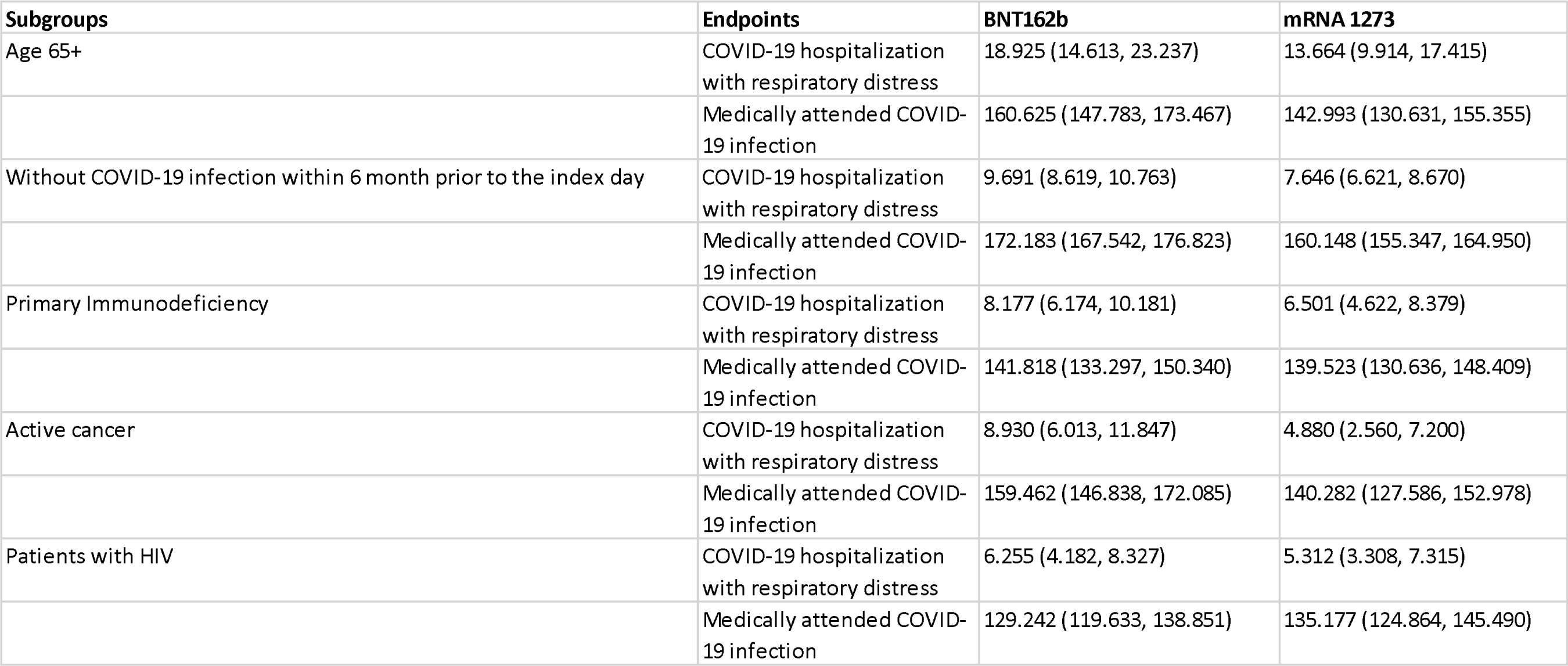
Crude rates of two study endpoints among subgroups.

| **Subgroups** | **Endpoints** | **BNT162b** | **mRNA 1273** |
| --- | --- | --- | --- |
| Age 65+ | COVID-19 hospitalization with respiratory distress | 18.925 (14.613, 23.237) | 13.664 (9.914, 17.415) |
|  | Medically attended COVID-19 infection | 160.625 (147.783, 173.467) | 142.993 (130.631, 155.355) |
| Without COVID-19 infection within 6 month prior to the index day | COVID-19 hospitalization with respiratory distress | 9.691 (8.619, 10.763) | 7.646 (6.621, 8.670) |
|  | Medically attended COVID-19 infection | 172.183 (167.542, 176.823) | 160.148 (155.347, 164.950) |
| Primary Immunodeficiency | COVID-19 hospitalization with respiratory distress | 8.177 (6.174, 10.181) | 6.501 (4.622, 8.379) |
|  | Medically attended COVID-19 infection | 141.818 (133.297, 150.340) | 139.523 (130.636, 148.409) |
| Active cancer | COVID-19 hospitalization with respiratory distress | 8.930 (6.013, 11.847) | 4.880 (2.560, 7.200) |
|  | Medically attended COVID-19 infection | 159.462 (146.838, 172.085) | 140.282 (127.586, 152.978) |
| Patients with HIV | COVID-19 hospitalization with respiratory distress | 6.255 (4.182, 8.327) | 5.312 (3.308, 7.315) |
|  | Medically attended COVID-19 infection | 129.242 (119.633, 138.851) | 135.177 (124.864, 145.490) |

**Supplemental Table 7.** The censoring reason distribution in main analysis.

|  | **BNT162b** | **mRNA 1273** | **Overall** |
| --- | --- | --- | --- |
| **COVID-19 hospitalization with respiratory distress** |  |  |  |
| Outcome | 342 (0.6%) | 232 (0.4%) | 574 (0.5%) |
| Specified date reached | 36,970 (61.5%) | 31,625 (59.7%) | 68,595 (60.7%) |
| End of patient enrollment | 10,928 (18.2%) | 9,191 (17.4%) | 20,119 (17.8%) |
| Receiving any COVID-19 vaccine after start of follow-up | 11,844 (19.7%) | 11,895 (22.5%) | 23,739 (21.0%) |
| **Medically attended COVID-19 infection** |  |  |  |
| Outcome | 5,784 (9.6%) | 4,656 (8.8%) | 10,440 (9.2%) |
| Specified date reached | 33,189 (55.2%) | 28,448 (53.7%) | 61,637 (54.5%) |
| End of patient enrollment | 10,021 (16.7%) | 8,608 (16.3%) | 18,629 (16.5%) |
| Receiving any COVID-19 vaccine after start of follow-up | 11,090 (18.5%) | 11,231 (21.2%) | 22,321 (19.7%) |

**Supplemental Figure 1.**
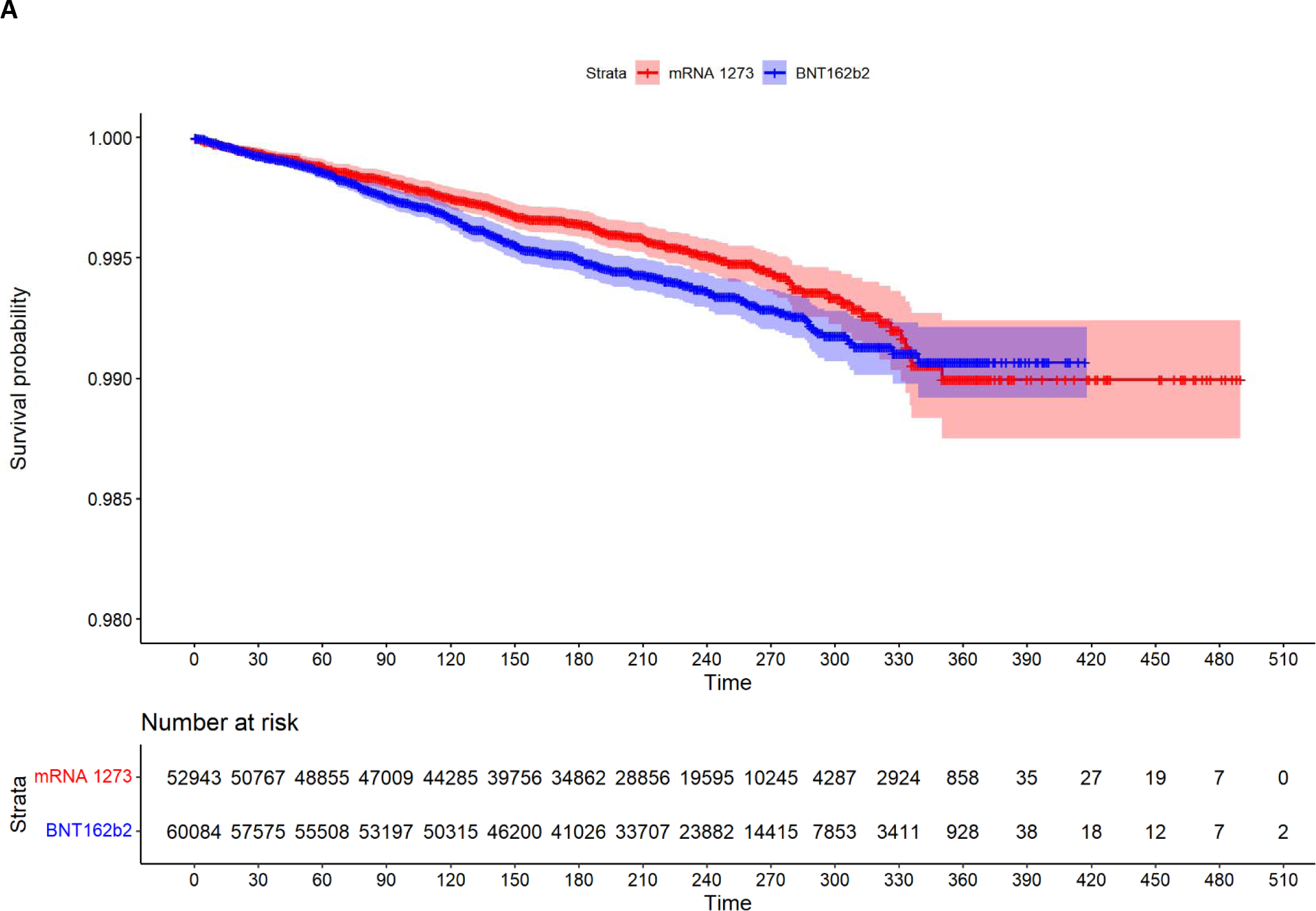
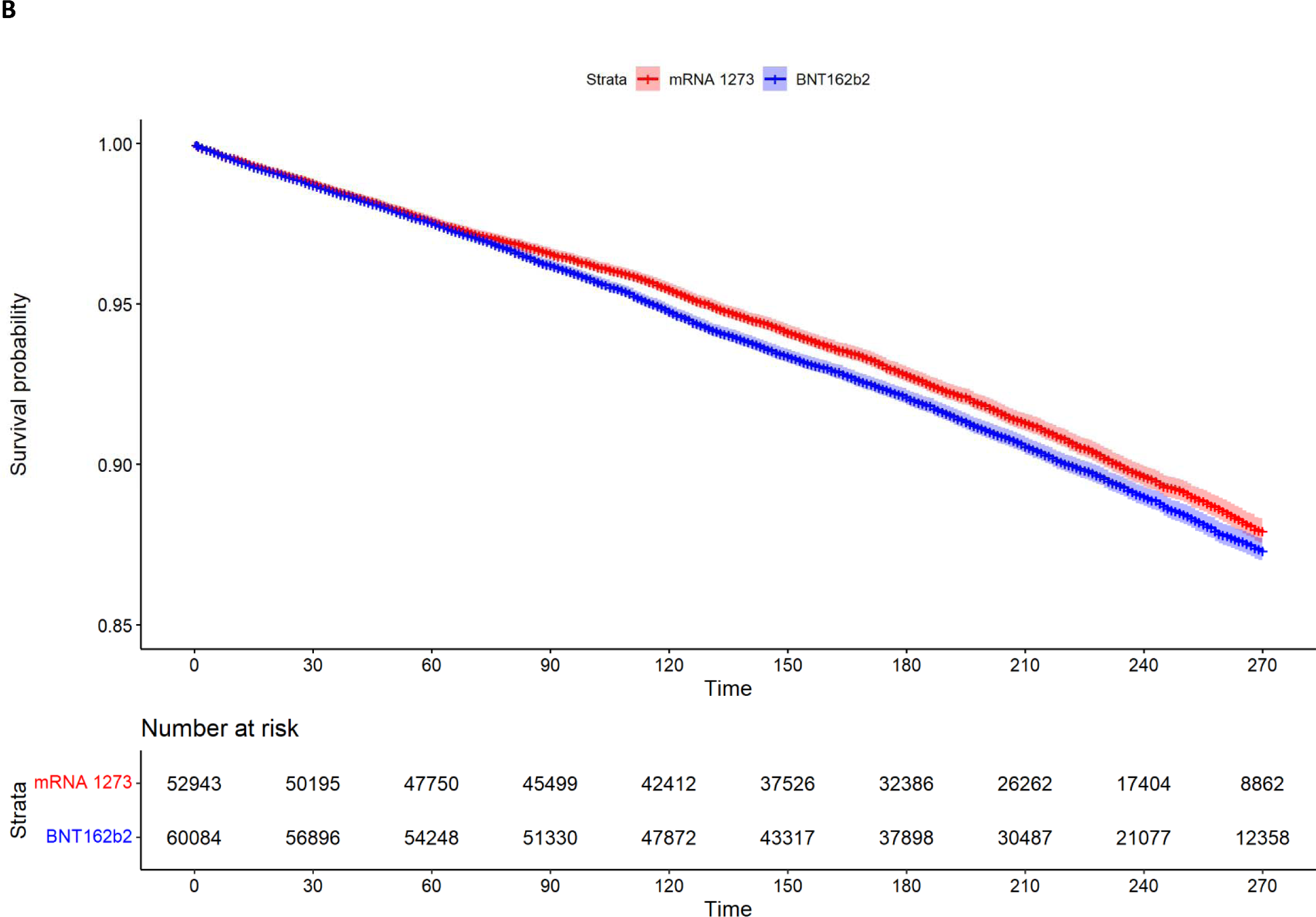
Kaplan–Meier plots of mRNA-1273 versus BNT162b cohort on A) COVID-19 hospitalization; B) Medically-attended COVID-19, throughout the full follow-up.

## REFERENCES

1. CDC. COVID Data Tracker. In: Centers for Disease and Control and Prevention, editor.: US Department of Health & Human Services; 2023

2. PAHO. Coronavirus disease (COVID-19). In: Pan American Health Organization, editor World Health Organization; 2023.

3. Ali K, Berman G, Zhou H, Deng W, Faughnan V, Coronado-Voges M, et al. Evaluation of mRNA-1273 SARS-CoV-2 Vaccine in Adolescents. N Engl J Med. 2021;385(24):2241–51.

4. Baden LR, El Sahly HM, Essink B, Kotloff K, Frey S, Novak R, et al. Efficacy and Safety of the mRNA-1273 SARS-CoV-2 Vaccine. N Engl J Med. 2021;384(5):403–16.

5. Polack FP, Thomas SJ, Kitchin N, Absalon J, Gurtman A, Lockhart S, et al. Safety and Efficacy of the BNT162b2 mRNA Covid-19 Vaccine. N Engl J Med. 2020;383(27):2603–15.

6. Bruxvoort KJ, Sy LS, Qian L, Ackerson BK, Luo Y, Lee GS, et al. Real-world effectiveness of the mRNA-1273 vaccine against COVID-19: Interim results from a prospective observational cohort study. Lancet Reg Health Am. 2022;6:100134.

7. Kow CS, Hasan SS. Real-world effectiveness of BNT162b2 mRNA vaccine: a meta-analysis of large observational studies. Inflammopharmacology. 2021;29(4):1075–90.

8. Feikin DR, Higdon MM, Abu-Raddad LJ, Andrews N, Araos R, Goldberg Y, et al. Duration of effectiveness of vaccines against SARS-CoV-2 infection and COVID-19 disease: results of a systematic review and meta-regression. Lancet. 2022;399(10328):924–44.

9. Tenforde MW, Patel MM, Gaglani M, Ginde AA, Douin DJ, Talbot HK, et al. Effectiveness of a Third Dose of Pfizer-BioNTech and Moderna Vaccines in Preventing COVID-19 Hospitalization Among Immunocompetent and Immunocompromised Adults - United States, August-December 2021. MMWR Morb Mortal Wkly Rep. 2022;71(4):118-24.

10. Moreira ED, Jr., Kitchin N, Xu X, Dychter SS, Lockhart S, Gurtman A, et al. Safety and Efficacy of a Third Dose of BNT162b2 Covid-19 Vaccine. N Engl J Med. 2022;386(20):1910–21.

11. Teo SP. Review of COVID-19 mRNA Vaccines: BNT162b2 and mRNA-1273. J Pharm Pract. 2022;35(6):947–51.

12. Lee A, Wong SY, Chai LYA, Lee SC, Lee MX, Muthiah MD, et al. Efficacy of covid-19 vaccines in immunocompromised patients: systematic review and meta-analysis. BMJ. 2022;376:e068632.

13. Chemaitelly H, Ayoub HH, Tang P, Coyle P, Yassine HM, Al Thani AA, et al. Long-term COVID-19 booster effectiveness by infection history and clinical vulnerability and immune imprinting: a retrospective population-based cohort study. Lancet Infect Dis. 2023;23(7):816–27.

14. Chemaitelly H, Tang P, Hasan MR, Al Mukdad S, Yassine HM, Benslimane FM, et al. Waning of BNT162b2 Vaccine Protection against SARS-CoV-2 Infection in Qatar. N Engl J Med. 2021;385(24):e83.

15. Munro APS, Janani L, Cornelius V, Aley PK, Babbage G, Baxter D, et al. Safety and immunogenicity of seven COVID-19 vaccines as a third dose (booster) following two doses of ChAdOx1 nCov-19 or BNT162b2 in the UK (COV-BOOST): a blinded, multicentre, randomised, controlled, phase 2 trial. Lancet. 2021;398(10318):2258–76.

16. Duly K, Farraye FA, Bhat S. COVID-19 vaccine use in immunocompromised patients: A commentary on evidence and recommendations. Am J Health Syst Pharm. 2022;79(2):63–71.

17. FDA. Factsheet for healthcare providers administering vaccine (vaccination providers) emergency use authorization (EUA) of the Moderna COVID-19 vaccine to prevent coronavirus disease 2019 (COVID-19). U.S. Food and Drug Administration.; 2022.

18. FDA. Vaccine Information Fact Sheet for Recipients and Caregivers About Comirnaty (COVID-19 Vaccine, mRNA) and Pfizer-BioNtech COVID-19 Vaccine to Prevent Coronavirus Disease 2019 (COVID-19). U.S. Food and Drug Administration. 2022.

19. Mues KE, Kirk B, Patel DA, Gelman A, Chavers LS, Talarico CA, et al. Real-world comparative effectiveness of mRNA-1273 and BNT162b2 vaccines among immunocompromised adults identified in administrative claims data in the United States. Vaccine. 2022;40(47):6730–9.

20. Ku JH, Sy LS, Qian L, Ackerson BK, Luo Y, Tubert JE, et al. Vaccine effectiveness of the mRNA-1273 3-dose primary series against COVID-19 in an immunocompromised population: A prospective observational cohort study. Vaccine. 2023;41(24):3636–46.

21. Florea A, Sy LS, Qian L, Ackerson BK, Luo Y, Tubert JE, et al. Effectiveness of Messenger RNA-1273 Vaccine Booster Against Coronavirus Disease 2019 in Immunocompetent Adults. Clin Infect Dis. 2023;76(2):252–62.

22. Barda N, Dagan N, Ben-Shlomo Y, Kepten E, Waxman J, Ohana R, et al. Safety of the BNT162b2 mRNA Covid-19 Vaccine in a Nationwide Setting. N Engl J Med. 2021; 385(12):1078–90.

23. Barda N, Dagan N, Cohen C, Hernán MA, Lipsitch M, Kohane IS, et al. Effectiveness of a third dose of the BNT162b2 mRNA COVID-19 vaccine for preventing severe outcomes in Israel: an observational study. Lancet. 2021;398(10316):2093–100.

24. Wang X, Haeussler K, Spellman A, Phillips LE, Ramiller A, Bausch-Jurken MT, et al. Comparative effectiveness of mRNA-1273 and BNT162b2 COVID-19 vaccines in immunocompromised individuals: a systematic review and meta-analysis using the GRADE framework. Front Immunol. 2023;14:1204831.

25. Stumpf J, Siepmann T, Lindner T, Karger C, Schwöbel J, Anders L, et al. Humoral and cellular immunity to SARS-CoV-2 vaccination in renal transplant versus dialysis patients: A prospective, multicenter observational study using mRNA-1273 or BNT162b2 mRNA vaccine. Lancet Reg Health Eur. 2021;9:100178.

26. Fernandes Q, Inchakalody VP, Merhi M, Mestiri S, Taib N, Moustafa Abo El-Ella D, et al. Emerging COVID-19 variants and their impact on SARS-CoV-2 diagnosis, therapeutics and vaccines. Ann Med. 2022;54(1):524–40.

27. CDC. Interim Clinical Considerations for Use of COVID-19 Vaccines in the United States. In: Prevention CfDCa, editor.: US Department of Health & Human Services; 2023.

28. Kluberg SA, Hou L, Dutcher SK, Billings M, Kit B, Toh S, et al. Validation of diagnosis codes to identify hospitalized COVID-19 patients in health care claims data. Pharmacoepidemiol Drug Saf. 2022;31(4):476–80.

29. Garry EM, Weckstein AR, Quinto K, et al. Use of an EHR to inform an administrative data algorithm to categorize inpatient COVID-19 severity. medRxiv 2021.10.04.21264513.

30. Quan H, Sundararajan V, Halfon P, Fong A, Burnand B, Luthi JC, et al. Coding algorithms for defining comorbidities in ICD-9-CM and ICD-10 administrative data. Med Care. 2005;43(11):1130–9.

31. Kim DH, Schneeweiss S, Glynn RJ, Lipsitz LA, Rockwood K, Avorn J. Measuring Frailty in Medicare Data: Development and Validation of a Claims-Based Frailty Index. J Gerontol A Biol Sci Med Sci. 2018;73(7):980–7.

32. Austin PC, Stuart EA. Moving towards best practice when using inverse probability of treatment weighting (IPTW) using the propensity score to estimate causal treatment effects in observational studies. Stat Med. 2015;34(28):3661–79.

33. Schulte PJ, Mascha EJ. Propensity Score Methods: Theory and Practice for Anesthesia Research. Anesth Analg. 2018;127(4):1074–84.

34. Wang SV, Verpillat P, Rassen JA, Patrick A, Garry EM, Bartels DB. Transparency and reproducibility of observational cohort studies using large healthcare databases. Clin Pharm Ther 2016;99:325–32

35. Chodick G, Tene L, Rotem RS, Patalon T, Gazit S, Ben-Tov A, et al. The Effectiveness of the Two-Dose BNT162b2 Vaccine: Analysis of Real-World Data. Clin Infect Dis. 2022;74(3): 472–8.

36. Hernán MA, Robins JM. Causal Inference: What If. Boca Raton: Chapman & Hall/CRC. https://www.hsph.harvard.edu/miguel-hernan/causal-inference-book, 2022.

37. Sashegyi A, Ferry D. On the interpretation of the hazard ratio and communication of survival benefit. Oncologist. 2017 Apr;22(4):484–486. doi: 10.1634/theoncologist.2016-0198. Epub 2017 Mar 17.

38. Oseran AS, Song Y, Xu J, et al. Long term risk of death and readmission after hospital admission with covid-19 among older adults: retrospective cohort study. BMJ. 2023; 382:e076222 doi:10.1136/bmj-2023-076222

39. Funk MJ, Landi SN. Misclassification in administrative claims data: quantifying the impact on treatment effect estimates. Curr Epidemiol Rep. 2014 Dec;1(4):175–185. doi: 10.1007/s40471-014-0027-z.

40. Bhatt AS, McElrath EE, Claggett BL, Bhatt DL, Adler DS, Solomon SD, Vaduganathan M. Accuracy of ICD-10 Diagnostic Codes to Identify COVID-19 Among Hospitalized Patients. J Gen Intern Med. 2021;36(8):2532–5.

41. Kadri SS, Gundrum J, Warner S, Cao Z, Babiker A, Klompas M, Rosenthal N. Uptake and Accuracy of the Diagnosis Code for COVID-19 Among US Hospitalizations. JAMA. 2020;324(24):2553–4.

42. Lynch KE, Viernes B, Gatsby E, DuVall SL, Jones BE, Box TL, et al. Positive Predictive Value of COVID-19 ICD-10 Diagnosis Codes Across Calendar Time and Clinical Setting. Clin Epidemiol. 2021;13:1011–8.

